# Safety of anti-thrombotic therapy in acute aortic dissection; single center, observational study

**DOI:** 10.1101/2022.05.18.22275251

**Authors:** Kensuke Hori, Nagisa Morikawa, Eiki Tayama, Yoshihiro Fukumoto

**Affiliations:** Division of Cardiovascular Medicine, Department of Internal Medicine, Kurume University School of Medicine; Department of Surgery, Kurume University School of Medicine

**Keywords:** acute aortic dissection, anti-thrombosis, mortality, anti-coagulant, anti-platelet

## Abstract

**Background:** Acute aortic dissection occurs due to a primary tear in aortic intima, with blood from aortic lumen penetrating into diseased media, in which anti-thrombotic therapies may be harmful. We examined the prognosis in patients, who had already taken antithrombotic therapies at the onset of acute aortic dissection, and the safety to administer anti-thrombotic drugs in acute phase during hospitalization.

**Methods and Results:** We retrospectively enrolled 685 patients with acute aortic dissection (type A/B: 454/231), who were transferred to Kurume University Hospital from 2004 to 2020. In both type A and B, there were no significant differences in in-hospital mortality between with and without antithrombotic therapies at the onset. Patients, who survived more than a day and administered anti-thrombotic drugs during hospitalization, had significantly lower in-hospital mortality than those who had no anti-thrombosis in acute phase in type A, while there was no significant difference in in-hospital mortality between the 2 groups in type B.

**Conclusions:** We demonstrated that anti-thrombotic drugs did not worsen the prognosis in patients with acute aortic dissection, indicating that we should not hesitate anti-thrombotic drugs if indicated.

## Introduction

Acute aortic dissection has poor prognosis,[1, 2] frequently resulting in sudden cardiac death. There are several classifications in acute aortic dissection, in which Stanford type A is classified as “all dissections involving the ascending aorta, regardless of the site of origin”, and Stanford type B is as “all dissections not involving the ascending aorta”.[3] Historically, the mortality of Stanford type A approaches 1% per hour,[4] and Stanford type B had 10 to 25% of mortality at 30 days.[3] Although diagnostic modalities and therapeutic strategies including surgical and endovascular treatment have recently advanced, mortality rate remains still high.[5]

Accumulated evidence has demonstrated that the inflammation plays a central role in the pathogenesis of acute aortic dissection.[6–8] Acute aortic dissection occurs due to a primary tear in aortic intima, with blood from aortic lumen penetrating into diseased media.[8] As initial medical therapy, blood pressure, heart rate, and pain should be initially treated to optimize patient stabilization during interhospital transfer and surgical triage.[9] In such situation, data regarding anti-thrombotic therapies remains scant, which seems to be harmful. However, acute aortic dissection suddenly occurs even in patients, who have already taken anti-thrombotic therapies, in the clinical settings. Further, anti-thrombotic drugs are occasionally required for several reasons, such as atrial fibrillation, during the hospitalization of acute aortic dissection.

Currently, it has not been fully elucidated if anti-thrombotic drugs do harm in patients with acute aortic dissection. Therefore, the present study aimed to examine the impact of anti-thrombotic therapies at the onset of acute aortic dissection and the safety of anti-thrombotic agents use during hospitalization in acute phase of acute aortic dissection.

## Methods

### Study Design

This study was a retrospective observational study using the database of Coronary Care Unit / Cardiovascular Medicine, Kurume University Hospital. We enrolled patients with acute aortic dissection, who were transferred to Coronary Care Unit / Cardiovascular Medicine, Kurume University Hospital from January 2004 to December 2020. The present study was approved by the institutional review board at Kurume University (21280). The informed consent was waived due to the retrospective nature and opt-out was used in the study.

### Data Collection

Baseline demographic data, when patients arrive at the hospital, were collected based on the medical records, including age, sex, height, body weight, waist, medications including anti-thrombosis (warfarin, direct oral anti-coagulants (DOAC), aspirin, clopidogrel), traditional risk factors (hypertension, glucose intolerance/diabetes mellitus and dyslipidemia), blood pressure (BP), pulse rate, heart rate, and comorbidities (coronary artery disease, hypertensive heart disease, cardiomyopathy, valvular heart diseases, and congenital heart diseases). Coronary artery diseases included chronic stable angina, asymptomatic myocardial ischemia, acute coronary syndrome, prior myocardial infarction, prior coronary revascularization, coronary spastic angina, and nonobstructive coronary atherosclerosis. Venous thromboembolism included pulmonary thromboembolism and deep vein thrombosis. Left ventricular (LV) dysfunction was defined as LV ejection fraction <50%. “Genetic and others” includes Marfan syndrome, Loeys-Dietz syndrome, and Behçet’s disease. Other clinical diagnosis and medication data were obtained from medical records. All patients were ascertained smoking and drinking habits by a questionnaire. Alcohol intake and smoking were classified as current habitual use or not. Major bleeding was defined as fatal bleeding and/or symptomatic bleeding in a critical area or organ, such as intracranial, intraspinal, intraocular, retroperitoneal, intraarticular or pericardial, or intramuscular with compartment syndrome, referred to International Society on Thrombosis and Haemostasis.[10] Infarction included myocardial, cerebral, and peripheral, including abdominal and limb, arteries infarction. Data of death during hospitalization were collected based on medical records. All cardiovascular diseases were diagnosed by expert cardiologists.

### Blood sampling

Blood samples were obtained from the antecubital vein and measured at a commercially available laboratory in Kurume University Hospital. Estimated glomerular filtration rate (eGFR) was calculated using the Modification of Diet in Renal Disease (MDRD) study equation modified with a Japanese coefficient[11]: eGFR (ml・min^-1^・1.73 m^-2^) = 194 × age^−0.287^ × serum creatinine^−1.094^ (if female×0.739).

### Outcome

Primary outcome was defined as death from any cause during hospitalization, all of which were identified in the medical records.

### Statistical Analysis

Data were presented mean ± standard error (SE) and compared between groups using two-sided *t*-tests. Categorical variables were presented as frequency and percent proportions and compared between groups using chi-square tests. Survival curves of death from all causes were estimated by the Kaplan-Meier method and compared using the log-rank test. Cox proportional hazards models were used to estimate hazard ratios (HRs) and 95% confidence intervals (CIs) for all-cause mortality. Next, we adjusted the models for age and sex. Statistical significance was defined as *p*<0.05. All statistical analyses were performed using the SAS system (Release 9.4, SAS Institute, Cary, NC, USA).

## Results

### Transferred patients

A total of 685 patients with acute aortic dissection were transferred to our hospital from January 2004 to December 2020 (**Figure 1**). There were 454 type A and 231 type B patients. In type A, 70 patients were treated by antithrombosis drugs on admission, including 24 with anti-coagulants, 39 anti-platelet drugs, and 7 both (**Figure 1**). Characteristics of transferred patients at baseline were shown in **Table 1**.

**Figure 1.**
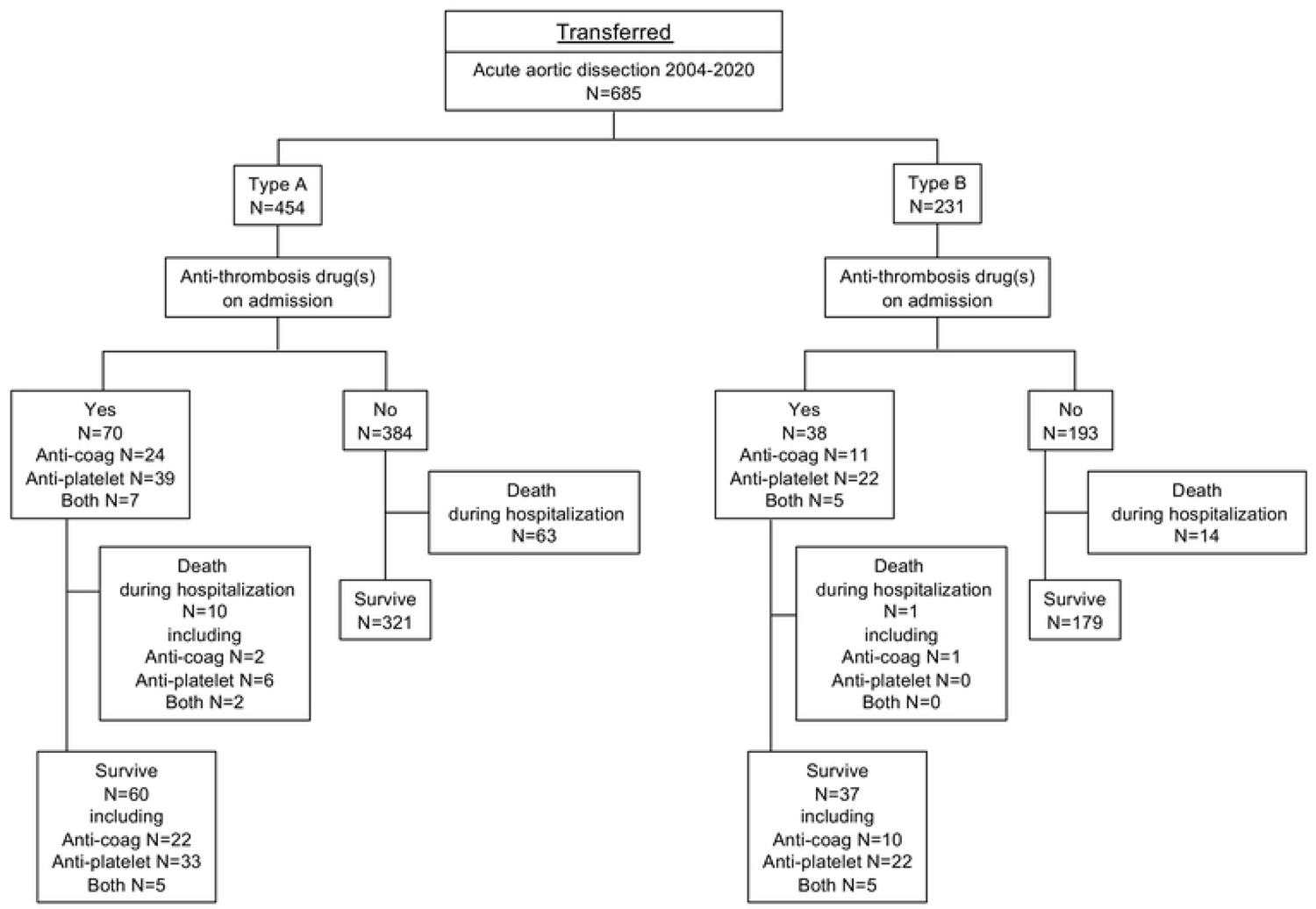
Enrollment of transferred patients with acute aortic dissection Among 685 transferred patients, 15% in 454 type A and 16% in 231 type B patients had already taken anti-thrombotic drugs.

**Table 1.**
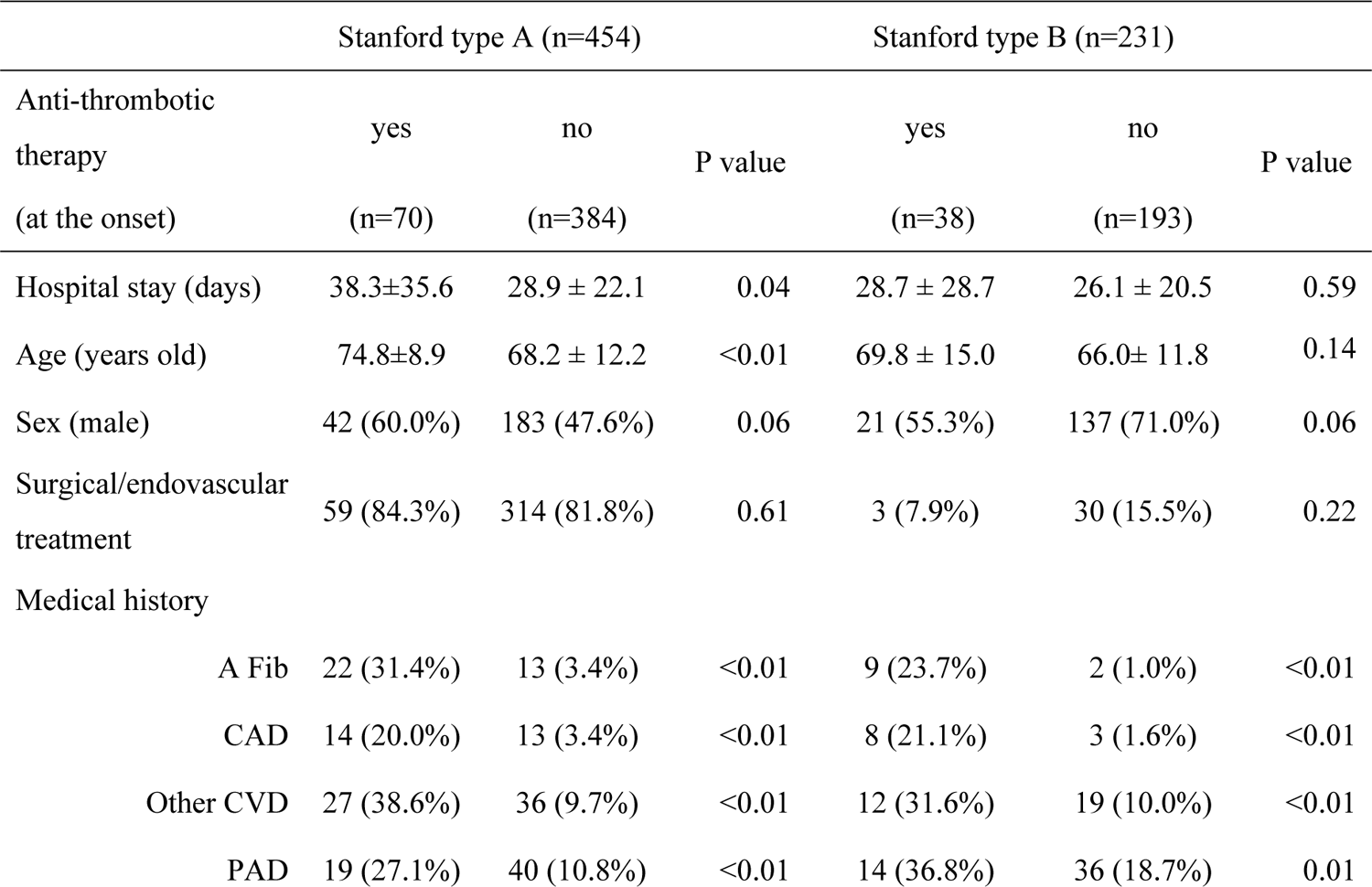
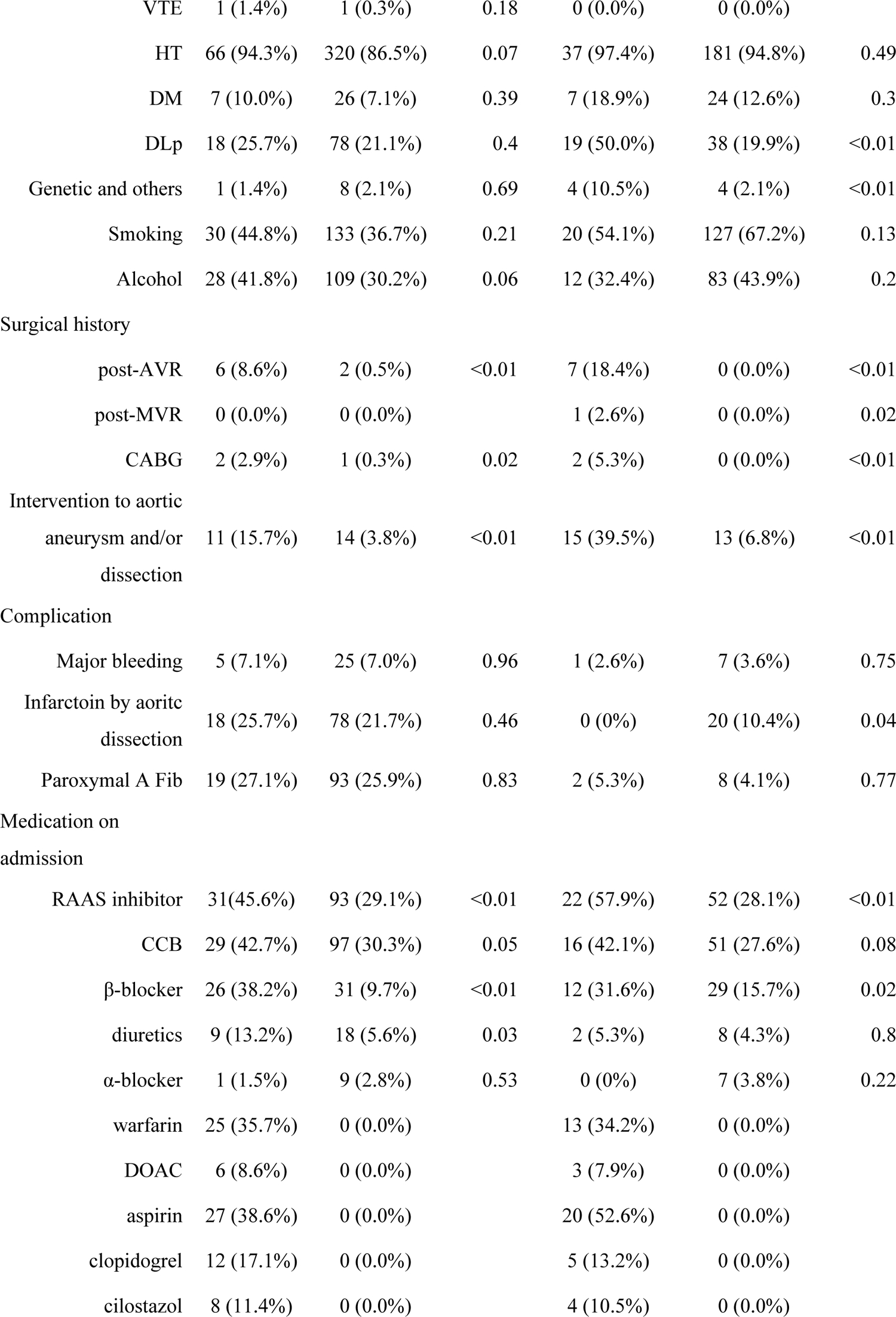
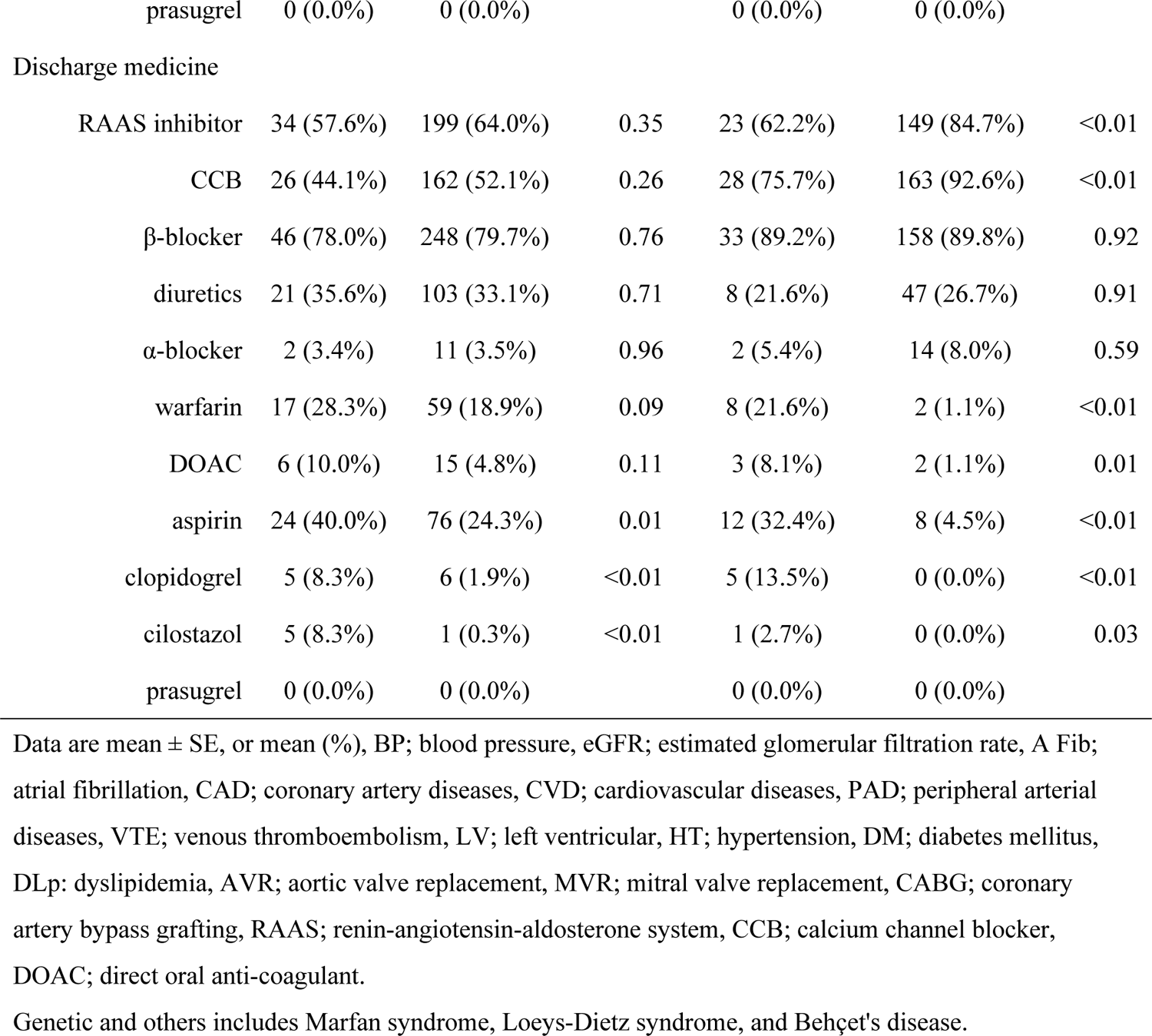
Clinical characteristics of transferred acute aortic dissection patients

In type A, patients, who were already treated by anti-thrombosis, had significantly older age, higher prevalence of previous history of atrial fibrillation, cardiovascular diseases, coronary artery bypass graft, and aortic valve replacement, and renin-angiotensin-aldosterone system (RAAS) inhibitors (angiotensin converting enzyme inhibitors and/or angiotensin II receptor blockers), calcium channel blocker, and β-blocker use. Among them, 10 patients (14.3%) died during the hospitalization (**Figure 1**).

In the remaining 384 patients with type A, who have not treated by anti-thrombosis drugs, 63 patients (16.4%) died during the hospitalization (**Figure 1**). In Kaplan-Meier curve, patients treated by antithrombosis drugs on admission did not show worse prognosis compared with no anti-thrombosis drug group in type A (**Figure 2A**).

**Figure 2.**
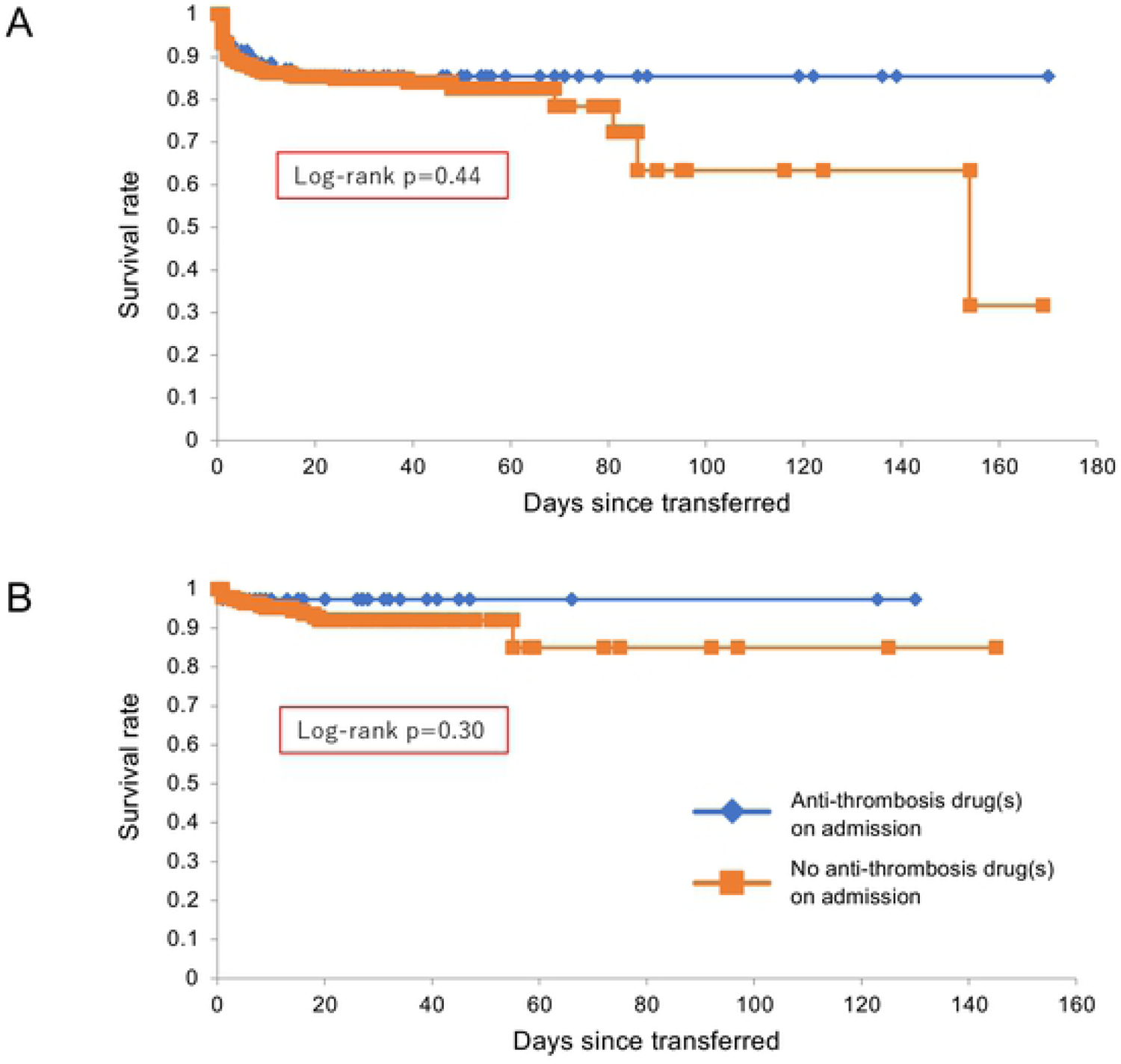
Kapan-Meier curves of transferred patients with acute aortic dissection. There were no significant differences in survival rate between with and without anti-thrombotic drugs on admission in both type A (panel A) and B (panel B).

In type B, 38 patients were treated by antithrombosis drugs on admission, including 11 with anti-coagulants, 22 anti-platelet drugs, and 5 both, who had significantly higher prevalence of previous history of atrial fibrillation, coronary artery disease, dyslipidemia, aortic valve replacement, and coronary artery bypass graft, and RAAS inhibitors use (**Figure 1**, **Table 1**). Among them, 1 patient (2.6%) died during the hospitalization. In the remaining 193 patients with type B, who have not treated by anti-thrombosis drugs, 14 patients (7.3%) died during the hospitalization. Kaplan-Meier curve indicated the comparable prognosis between the 2 groups of type B (**Figure 2B**).

### Hospitalized patients

A total of 650 patients with acute aortic dissection survived for more than a day and hospitalized (**Figure 3**). There were 424 type A and 226 type B patients. In type A, 182 patients were treated by antithrombosis drugs during hospitalization, including 64 with anti-coagulants, 73 anti-platelet drugs, and 45 both. Characteristics of hospitalized patients at baseline were shown in **Table 2**. Type A patients had significantly higher prevalence of atrial fibrillation and genetic and other aortic diseases, including Marfan syndrome, Loeys-Dietz syndrome, and Behçet’s disease (**Table 2**).

**Figure 3.**
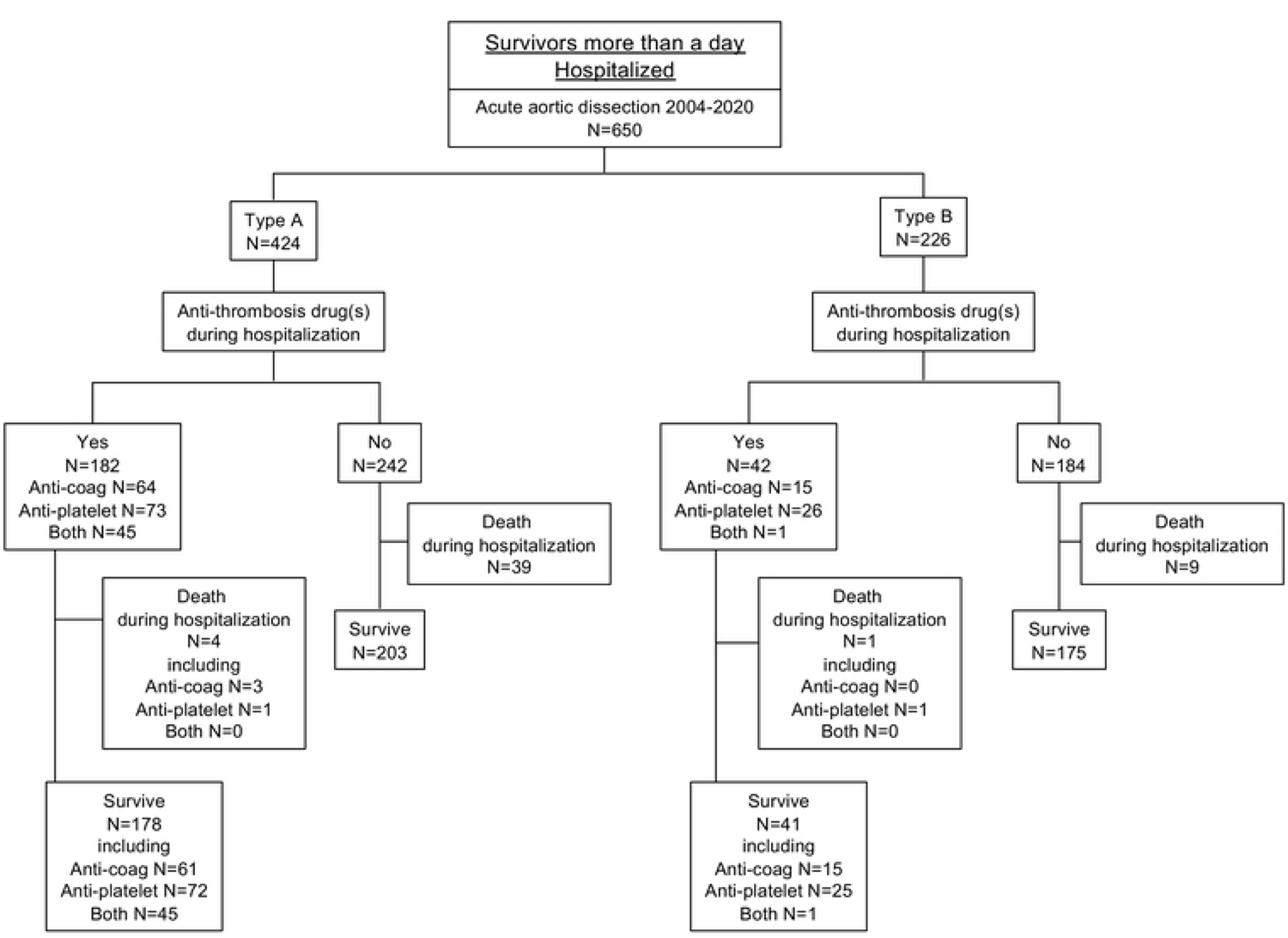
Enrollment of hospitalized patients with acute aortic dissection Among 650 hospitalized patients, 43% in 424 type A and 19% in 225 type B patients were treated by anti-thrombotic drugs during hospitalization.

**Table 2.**
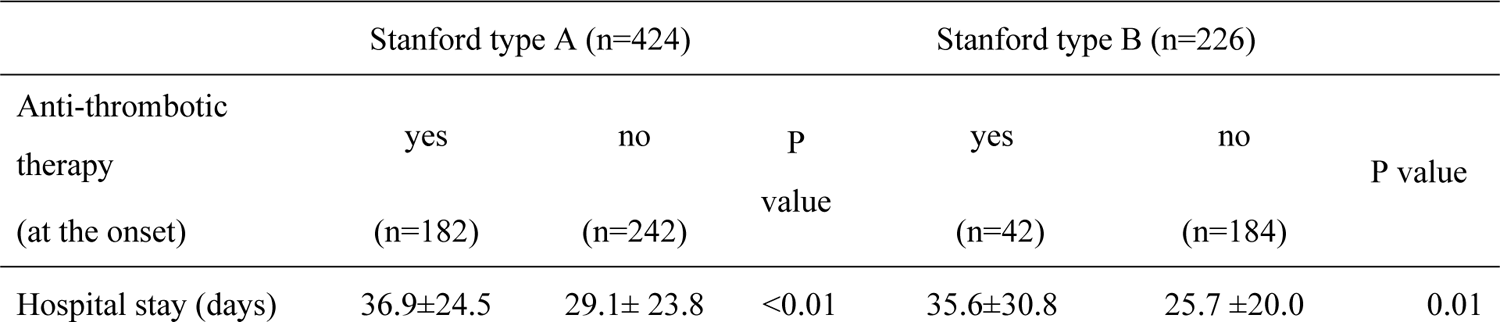
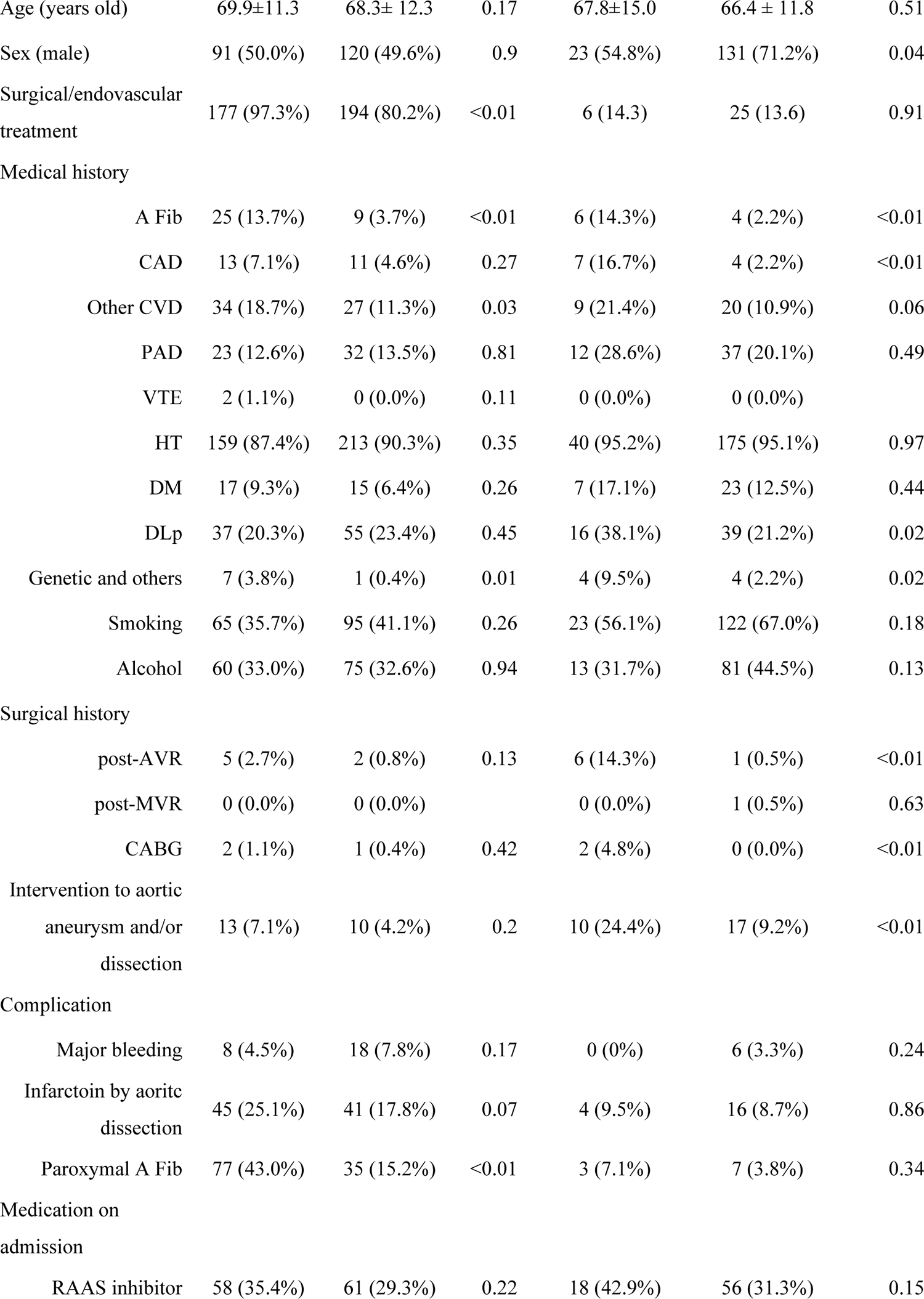
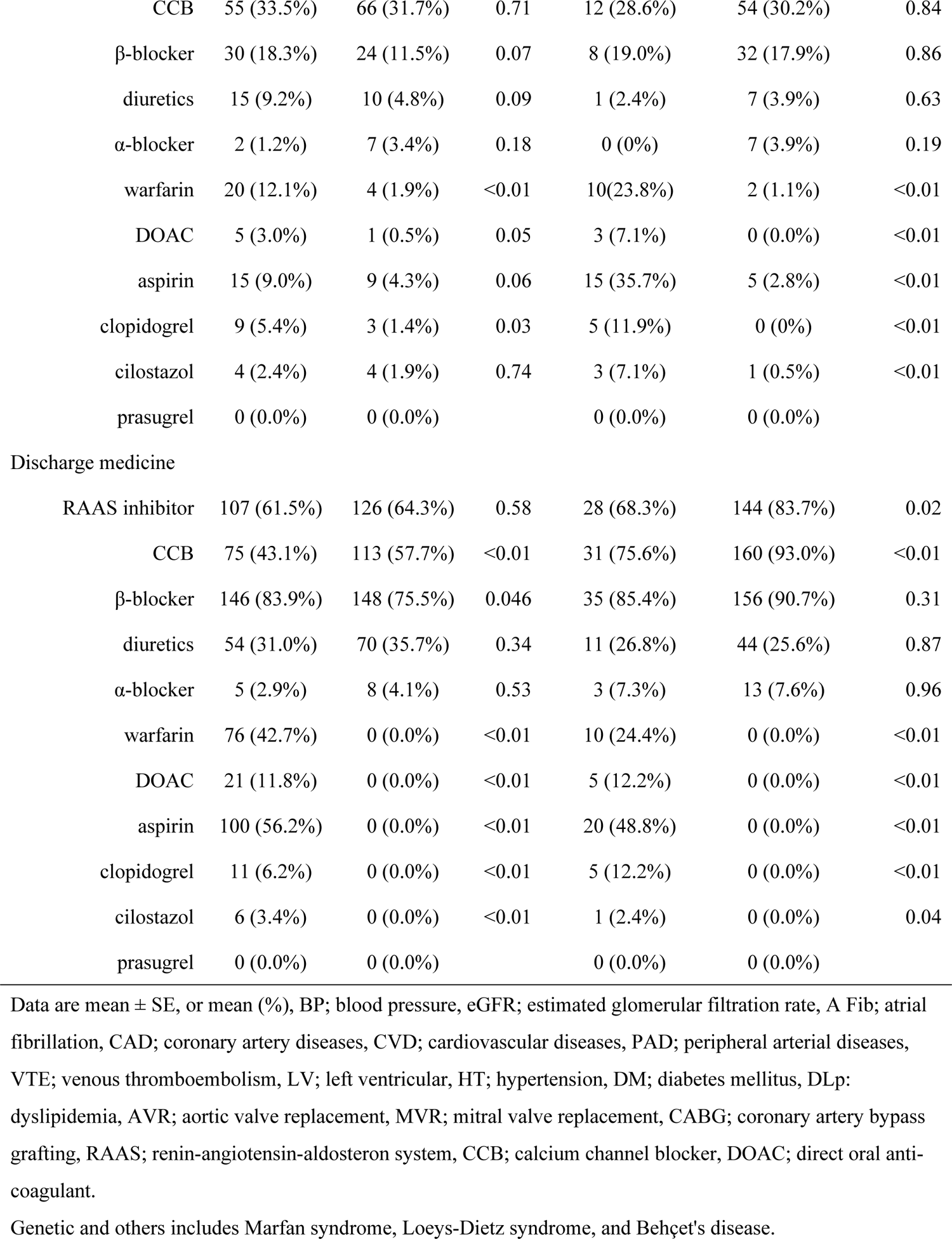
Clinical characteristics of hospitalized acute aortic dissection

Among them, 4 patients (2.2%) died during hospitalization, and 178 (97.8% were discharged (**Figure 3**). In the remaining 242 patients with type A, who have not been treated by anti-thrombosis drugs, 39 patients (16.1%) died and 203 (83.9%) were discharged (**Figure 3**). Kaplan-Meier curve showed the significantly better prognosis in patients treated by antithrombosis drugs in type A (**Figure 4A**).

**Figure 4.**
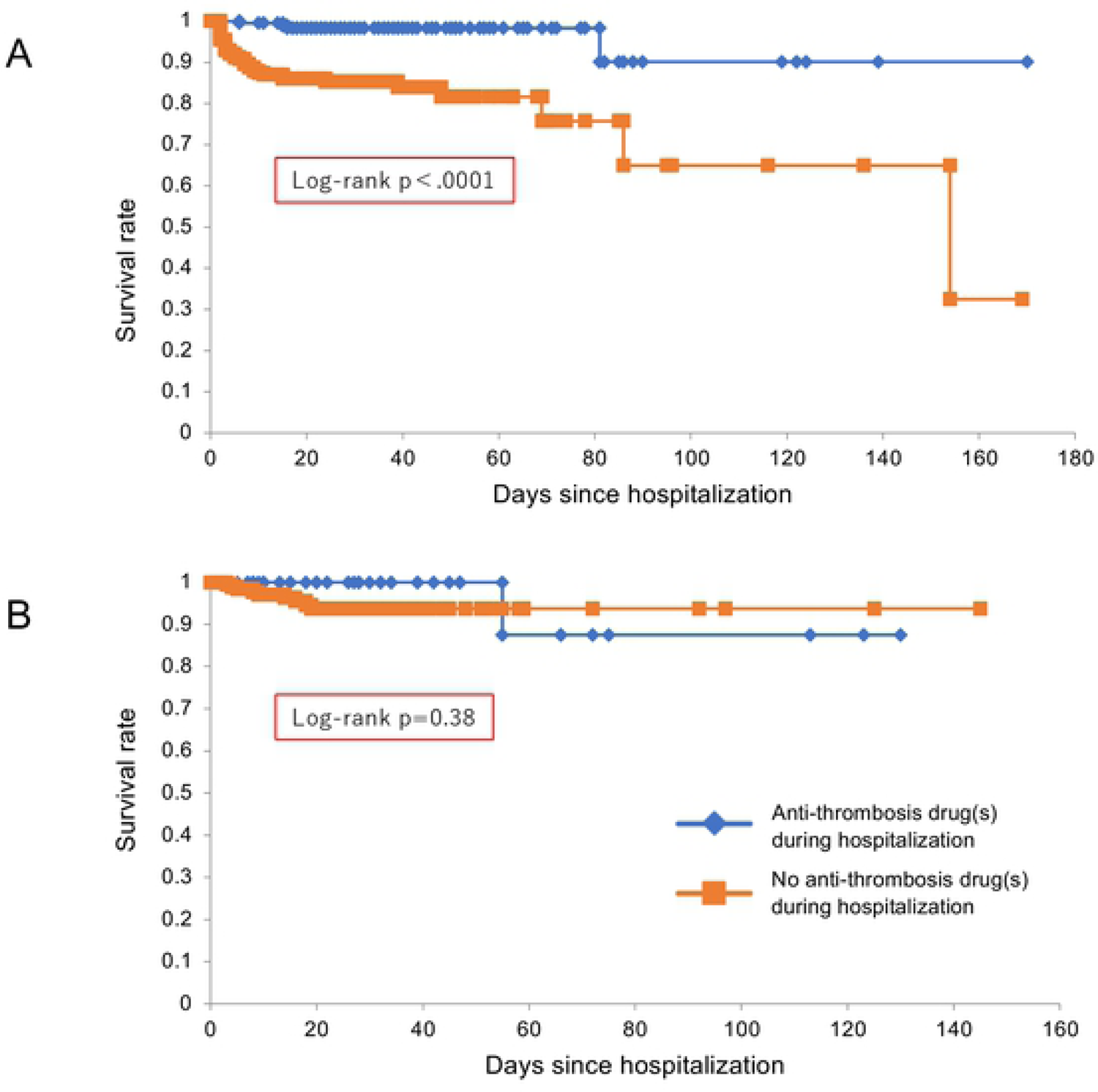
Kapan-Meier curves of hospitalized patients with acute aortic dissection. Survival rate was significantly better in anti-thrombotic drug group than the other in type A (panel A), while there was no significant difference between with and without anti-thrombotic drugs during hospitalization in type B (panel B).

In 226 type B patients, 42 patients were treated by antithrombosis drugs, including 15 with anti-coagulants, 26 anti-platelet drugs, and 1 both (**Figure 3**), who had significantly higher prevalence of atrial fibrillation, coronary artery diseases, aortic valve replacement, and coronary artery bypass graft (**Table 2**). Among them, 1 patient (2.4%) died, and 41 were discharged. In the remaining 184 type B patients, who have not treated by anti-thrombosis drugs, 9 patients (4.9%) died during hospitalization (**Figure 3**). In Kaplan-Meier curve, there were no significant difference in prognosis in type B (**Figure 4B**).

### Cox proportional hazards regression analysis of all-cause death in TRANSFERRED patients in type A

In the Cox proportional hazards regression analysis of all-cause death, systolic blood pressure, eGFR, history of hypertension, smoking habit, alcohol intake, complication of surgical procedures, and use of α-blocker were significantly associated with all-cause death (**Table 3**). In this analysis, the use of anti-thrombosis (warfarin, DOAC, aspirin, clopidogrel) was not the predictor of all cause death (**Table 3**).

**Table 3.**
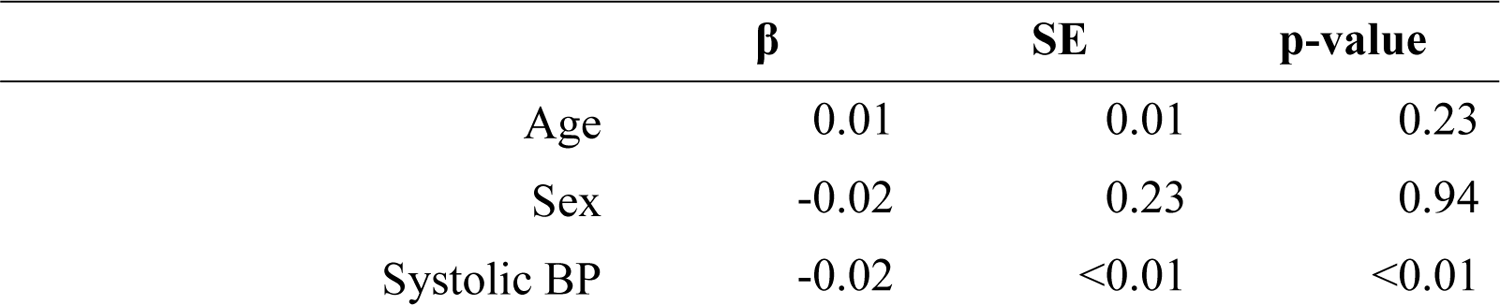
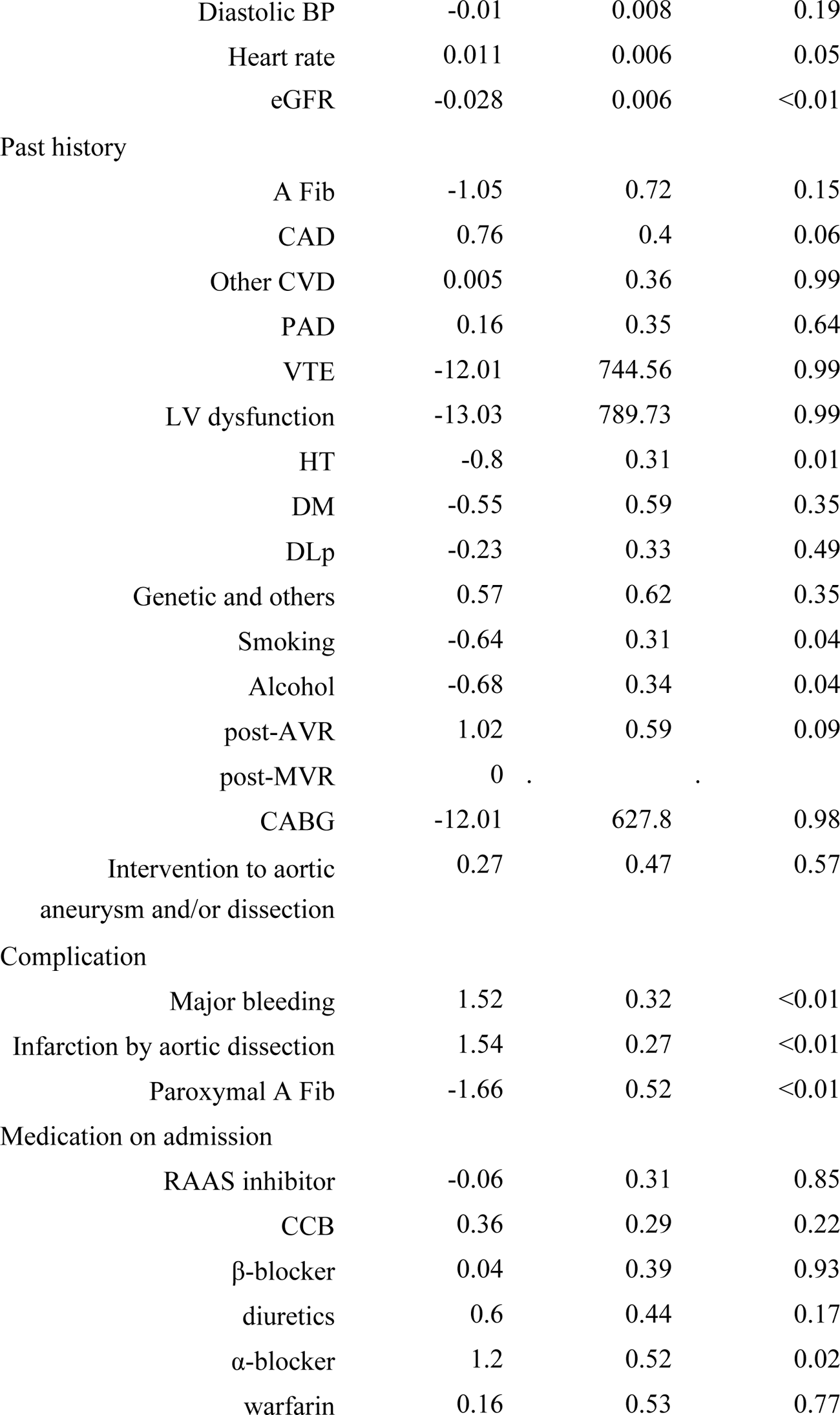
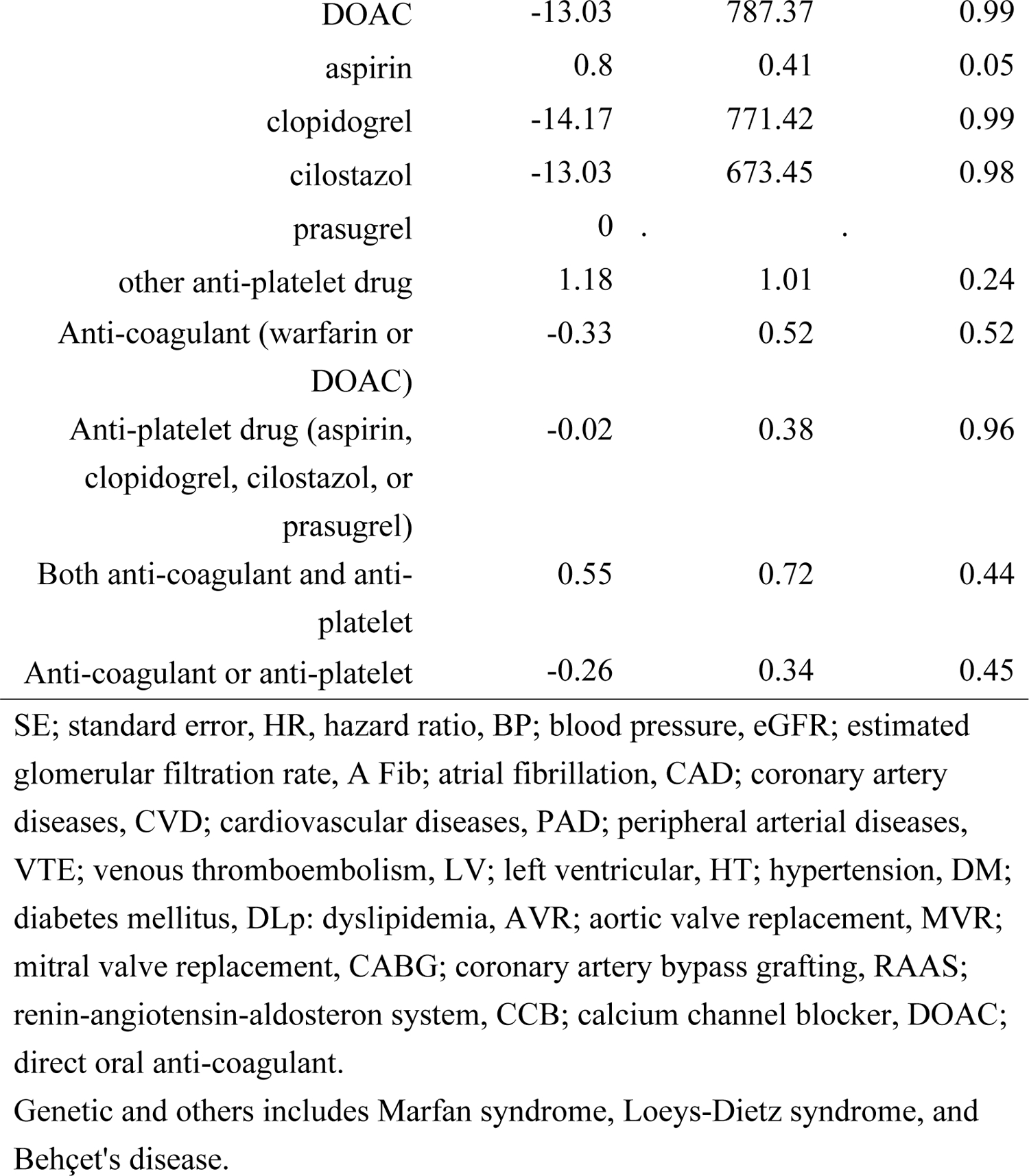
Cox proportional hazard model in transferred acute aortic dissection patients (type A)

| | $\beta$ | SE | p-value |
| --- | --- | --- | --- |
| Age | 0.01 | 0.01 | 0.23 |
| Sex | -0.02 | 0.23 | 0.94 |
| Systolic BP | -0.02 | <0.01 | <0.01 |

|  |  |  |  |  |
| --- | --- | --- | --- | --- |
|  | Diastolic BP | -0.01 | 0.008 | 0.19 |
|  | Heart rate | 0.011 | 0.006 | 0.05 |
|  | eGFR | -0.028 | 0.006 | <0.01 |
| Past history |  |  |  |  |
|  | A Fib | -1.05 | 0.72 | 0.15 |
|  | CAD | 0.76 | 0.4 | 0.06 |
|  | Other CVD | 0.005 | 0.36 | 0.99 |
|  | PAD | 0.16 | 0.35 | 0.64 |
|  | VTE | -12.01 | 744.56 | 0.99 |
|  | LV dysfunction | -13.03 | 789.73 | 0.99 |
|  | HT | -0.8 | 0.31 | 0.01 |
|  | DM | -0.55 | 0.59 | 0.35 |
|  | DLp | -0.23 | 0.33 | 0.49 |
|  | Genetic and others | 0.57 | 0.62 | 0.35 |
|  | Smoking | -0.64 | 0.31 | 0.04 |
|  | Alcohol | -0.68 | 0.34 | 0.04 |
|  | post-AVR | 1.02 | 0.59 | 0.09 |
|  | post-MVR | 0 |  |  |
|  | CABG | -12.01 | 627.8 | 0.98 |
|  | Intervention to aortic aneurysm and/or dissection | 0.27 | 0.47 | 0.57 |
| Complication |  |  |  |  |
|  | Major bleeding | 1.52 | 0.32 | <0.01 |
|  | Infarction by aortic dissection | 1.54 | 0.27 | <0.01 |
|  | Paroxymal A Fib | -1.66 | 0.52 | <0.01 |
| Medication on admission |  |  |  |  |
|  | RAAS inhibitor | -0.06 | 0.31 | 0.85 |
|  | CCB | 0.36 | 0.29 | 0.22 |
| | β-blocker | 0.04 | 0.39 | 0.93 |
|  | diuretics | 0.6 | 0.44 | 0.17 |
| | α-blocker | 1.2 | 0.52 | 0.02 |
|  | warfarin | 0.16 | 0.53 | 0.77 |
| DOAC | -13.03 | 787.37 | 0.99 |
| aspirin | 0.8 | 0.41 | 0.05 |
| clopidogrel | -14.17 | 771.42 | 0.99 |
| cilostazol | -13.03 | 673.45 | 0.98 |
| prasugrel | 0 | . | . |
| other anti-platelet drug | 1.18 | 1.01 | 0.24 |
| Anti-coagulant (warfarin or DOAC) | -0.33 | 0.52 | 0.52 |
| Anti-platelet drug (aspirin, clopidogrel, cilostazol, or prasugrel) | -0.02 | 0.38 | 0.96 |
| Both anti-coagulant and anti-platelet | 0.55 | 0.72 | 0.44 |
| Anti-coagulant or anti-platelet | -0.26 | 0.34 | 0.45 |

SE; standard error, HR, hazard ratio, BP; blood pressure, eGFR; estimated glomerular filtration rate, A Fib; atrial fibrillation, CAD; coronary artery diseases, CVD; cardiovascular diseases, PAD; peripheral arterial diseases, VTE; venous thromboembolism, LV; left ventricular, HT; hypertension, DM; diabetes mellitus, DLp: dyslipidemia, AVR; aortic valve replacement, MVR; mitral valve replacement, CABG; coronary artery bypass grafting, RAAS; renin-angiotensin-aldosterone system, CCB; calcium channel blocker, DOAC; direct oral anti-coagulant.
Genetic and others includes Marfan syndrome, Loeys-Dietz syndrome, and Behçet's disease.

After adjustment for age and sex, systolic blood pressure, eGFR, history of hypertension, smoking habit, alcohol intake, complication of surgical procedures, and use of α-blocker were significantly associated with all-cause death (**Table 4**). Also, the use of anti-thrombosis (warfarin, DOAC, aspirin, clopidogrel) was not the predictor of all cause death (**Table 4**).

**Table 4.**
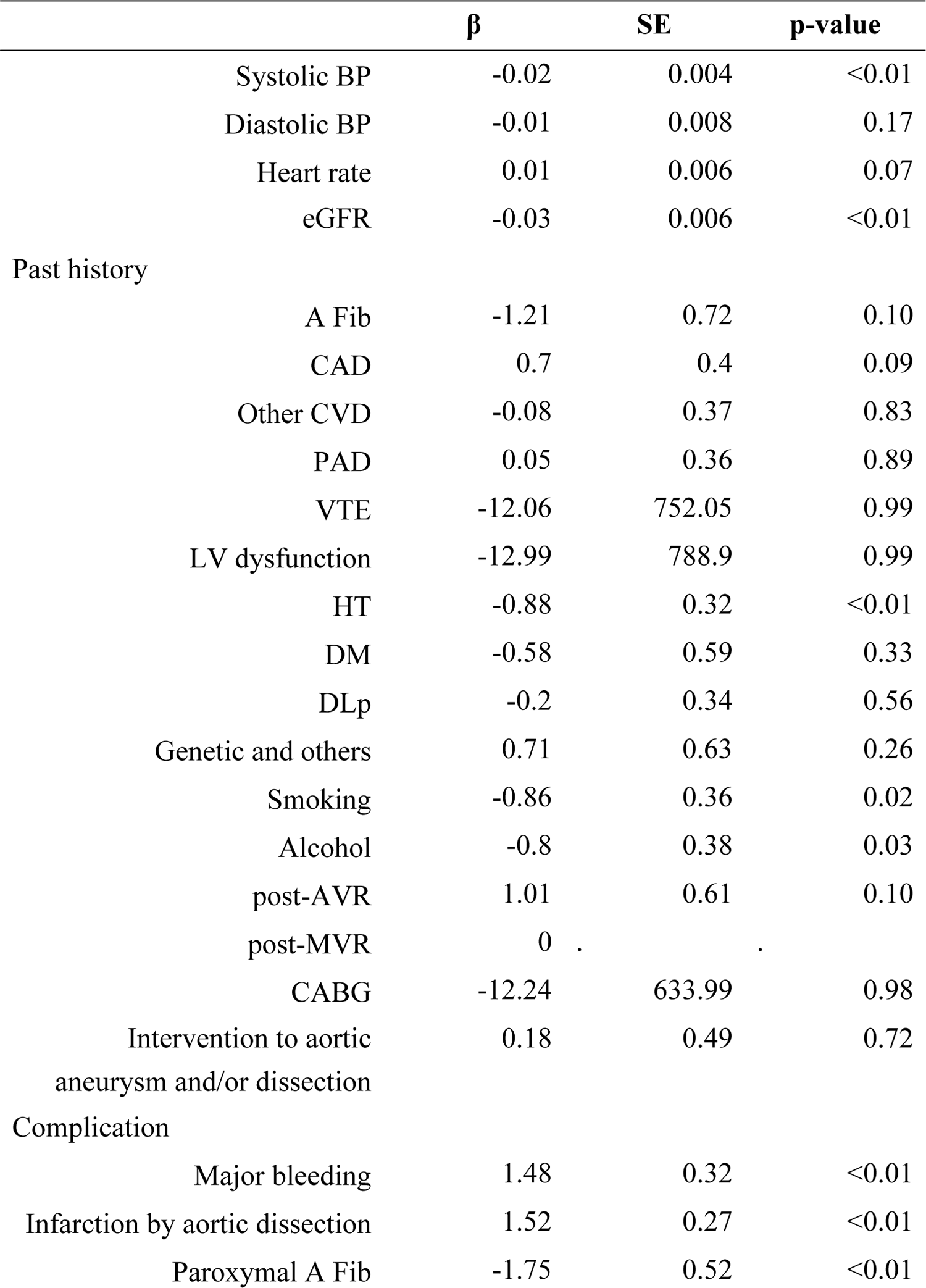
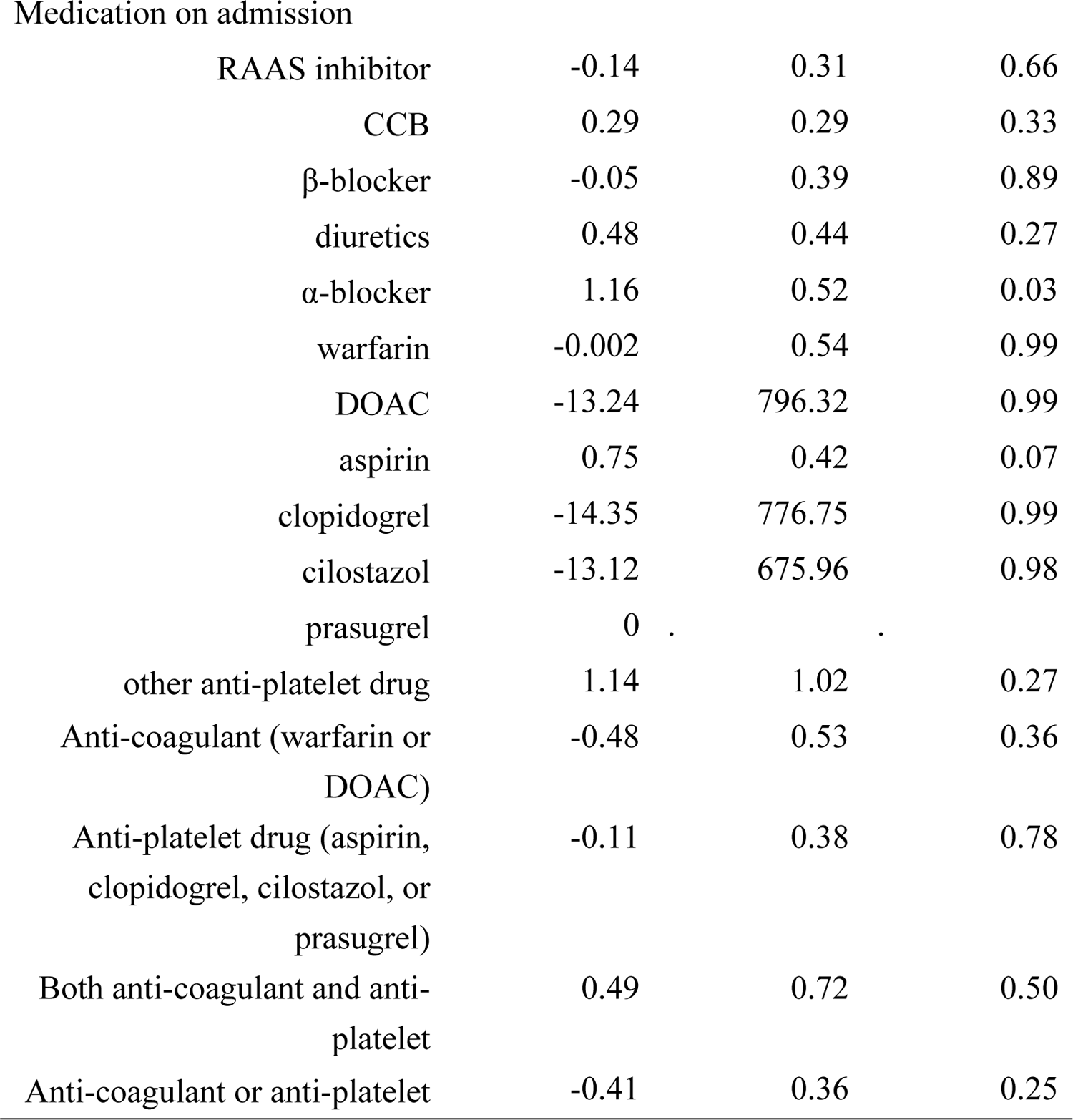
Age and sex-adjusted Cox proportional hazard model in transferred acute aortic dissection patients (type A)

| | $\beta$ | SE | p-value |
| --- | --- | --- | --- |
| Systolic BP | -0.02 | 0.004 | <0.01 |
| Diastolic BP | -0.01 | 0.008 | 0.17 |
| Heart rate | 0.01 | 0.006 | 0.07 |
| eGFR | -0.03 | 0.006 | <0.01 |
| Past history |  |  |  |
| A Fib | -1.21 | 0.72 | 0.10 |
| CAD | 0.7 | 0.4 | 0.09 |
| Other CVD | -0.08 | 0.37 | 0.83 |
| PAD | 0.05 | 0.36 | 0.89 |
| VTE | -12.06 | 752.05 | 0.99 |
| LV dysfunction | -12.99 | 788.9 | 0.99 |
| HT | -0.88 | 0.32 | <0.01 |
| DM | -0.58 | 0.59 | 0.33 |
| DLp | -0.2 | 0.34 | 0.56 |
| Genetic and others | 0.71 | 0.63 | 0.26 |
| Smoking | -0.86 | 0.36 | 0.02 |
| Alcohol | -0.8 | 0.38 | 0.03 |
| post-AVR | 1.01 | 0.61 | 0.10 |
| post-MVR | 0 |  |  |
| CABG | -12.24 | 633.99 | 0.98 |
| Intervention to aortic<br>aneurysm and/or dissection | 0.18 | 0.49 | 0.72 |
| Complication |  |  |  |
| Major bleeding | 1.48 | 0.32 | <0.01 |
| Infarction by aortic dissection | 1.52 | 0.27 | <0.01 |
| Paroxymal A Fib | -1.75 | 0.52 | <0.01 |
| Medication on admission |  |  |  |
| RAAS inhibitor | -0.14 | 0.31 | 0.66 |
| CCB | 0.29 | 0.29 | 0.33 |
| β-blocker | -0.05 | 0.39 | 0.89 |
| diuretics | 0.48 | 0.44 | 0.27 |
| α-blocker | 1.16 | 0.52 | 0.03 |
| warfarin | -0.002 | 0.54 | 0.99 |
| DOAC | -13.24 | 796.32 | 0.99 |
| aspirin | 0.75 | 0.42 | 0.07 |
| clopidogrel | -14.35 | 776.75 | 0.99 |
| cilostazol | -13.12 | 675.96 | 0.98 |
| prasugrel | 0 | . | . |
| other anti-platelet drug | 1.14 | 1.02 | 0.27 |
| Anti-coagulant (warfarin or DOAC) | -0.48 | 0.53 | 0.36 |
| Anti-platelet drug (aspirin, clopidogrel, cilostazol, or prasugrel) | -0.11 | 0.38 | 0.78 |
| Both anti-coagulant and anti-platelet | 0.49 | 0.72 | 0.50 |
| Anti-coagulant or anti-platelet | -0.41 | 0.36 | 0.25 |

### Cox proportional hazards regression analysis of all-cause death in TRANSFERRED patients in type B

In the Cox proportional hazards regression analysis of all-cause death, systolic blood pressure, heart rate, eGFR, the history of atrial fibrillation, complication of surgical procedures, and use of diuretics were significantly associated with all-cause death (**Table 5**). Also in this analysis, the use of anti-thrombosis (warfarin, DOAC, aspirin, clopidogrel) was not the predictor of all cause death (**Table 5**).

**Table 5.**
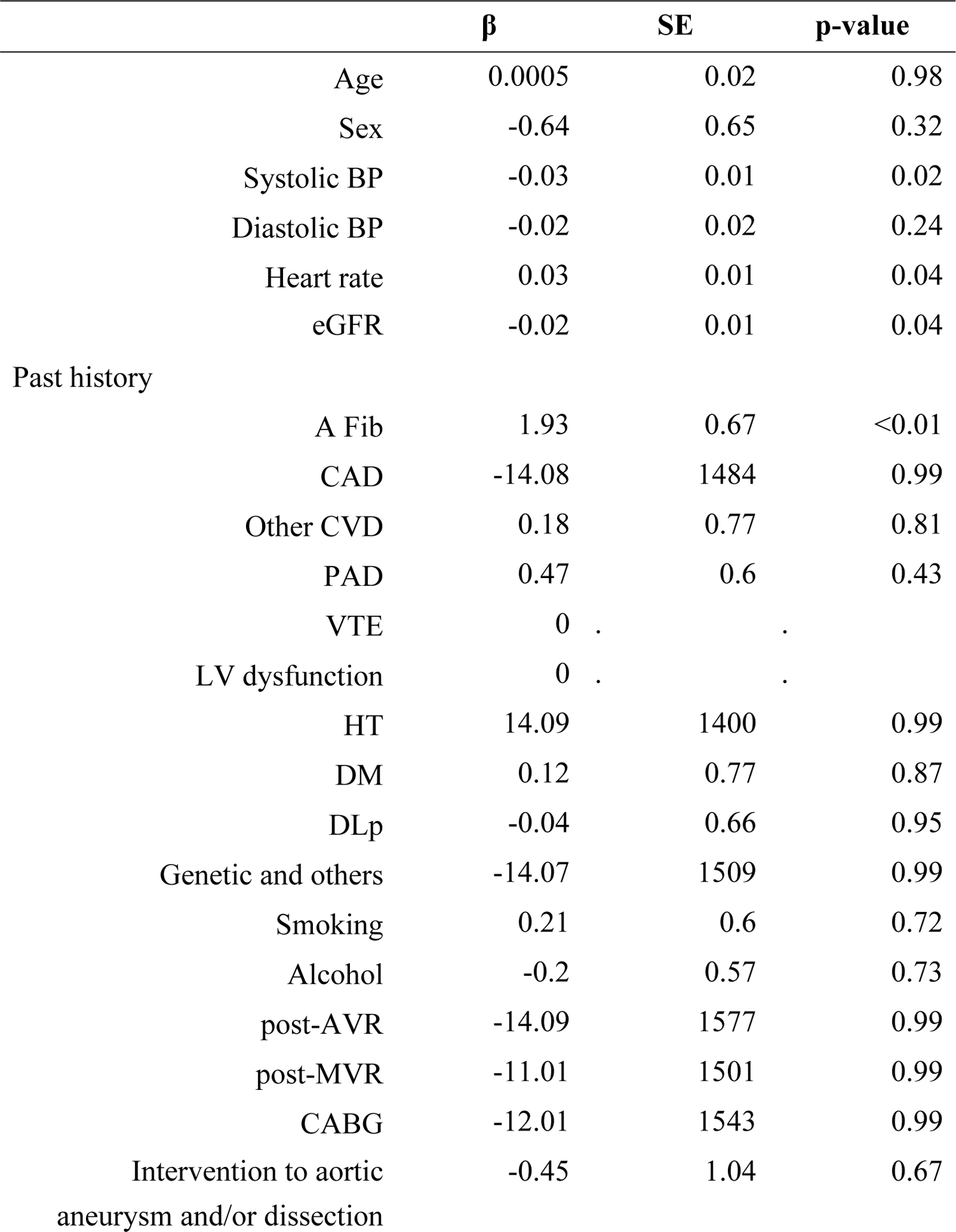
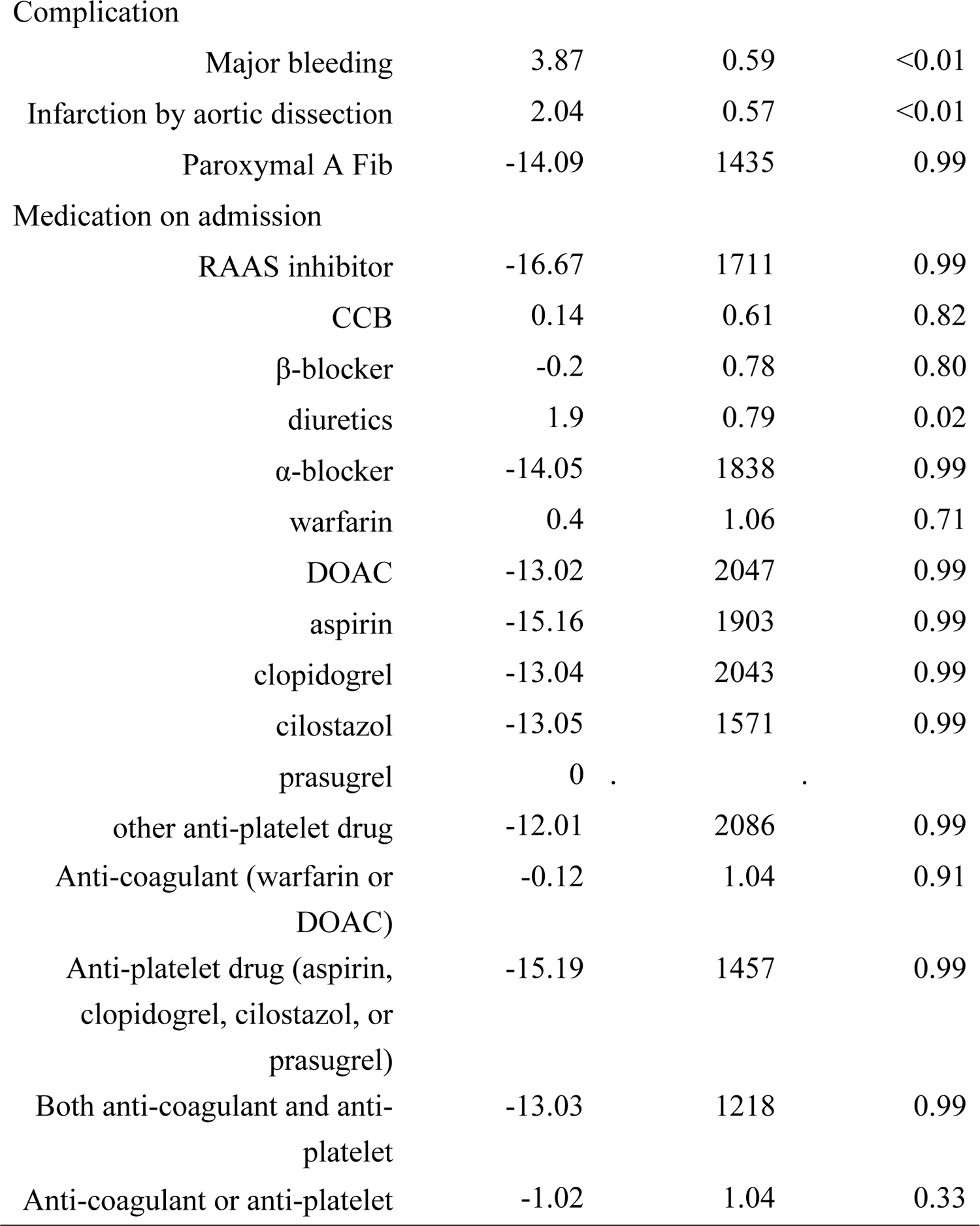
Cox proportional hazard model in transferred acute aortic dissection patients (type B)

After adjustment for age and sex, systolic blood pressure, heart rate, eGFR, the history of atrial fibrillation, complication of surgical procedures, and use of diuretics were significantly associated with all-cause death (**Table 6**), and the use of anti-thrombosis was not the predictor of all cause death (**Table 6**).

**Table 6.**
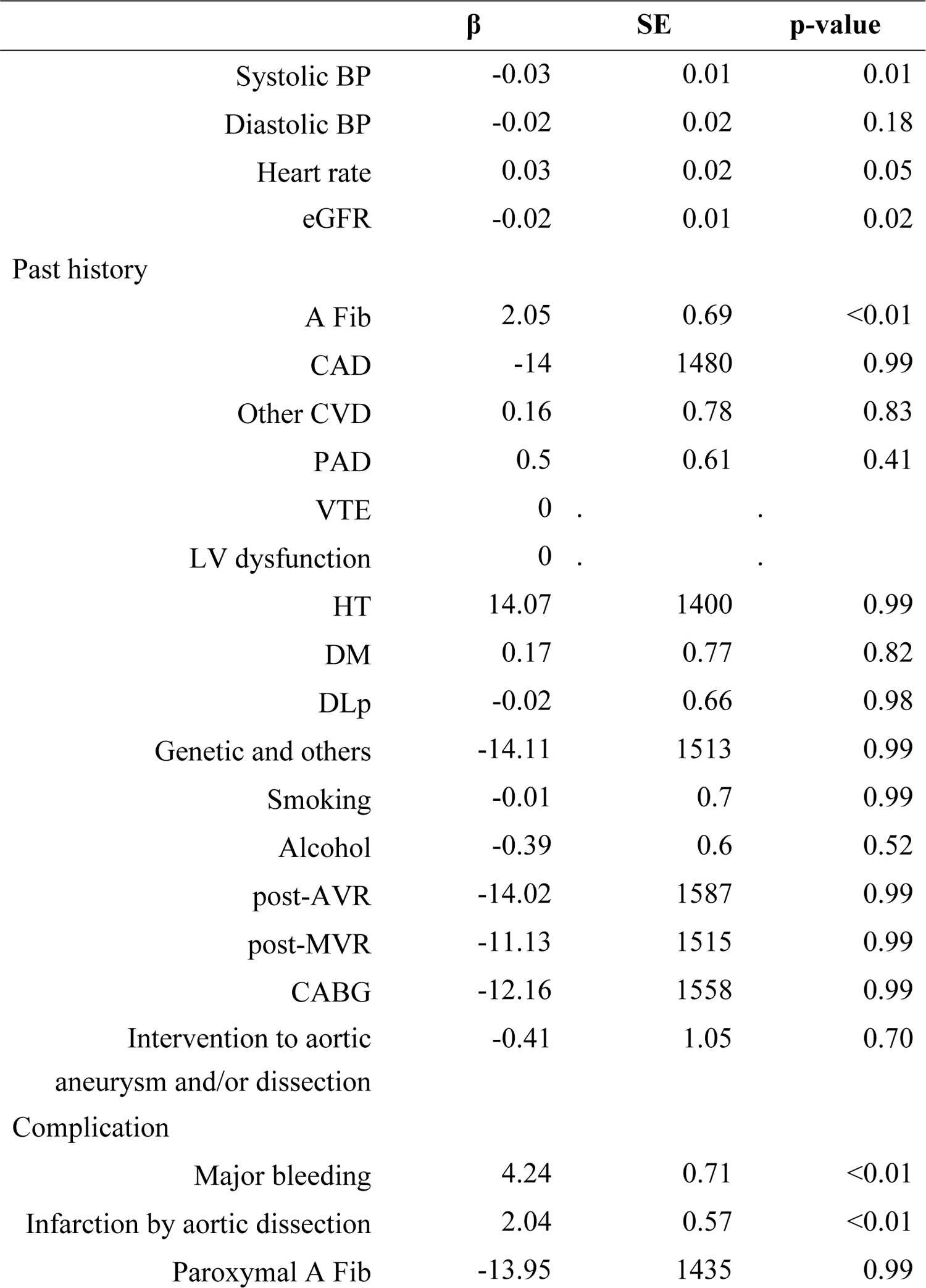
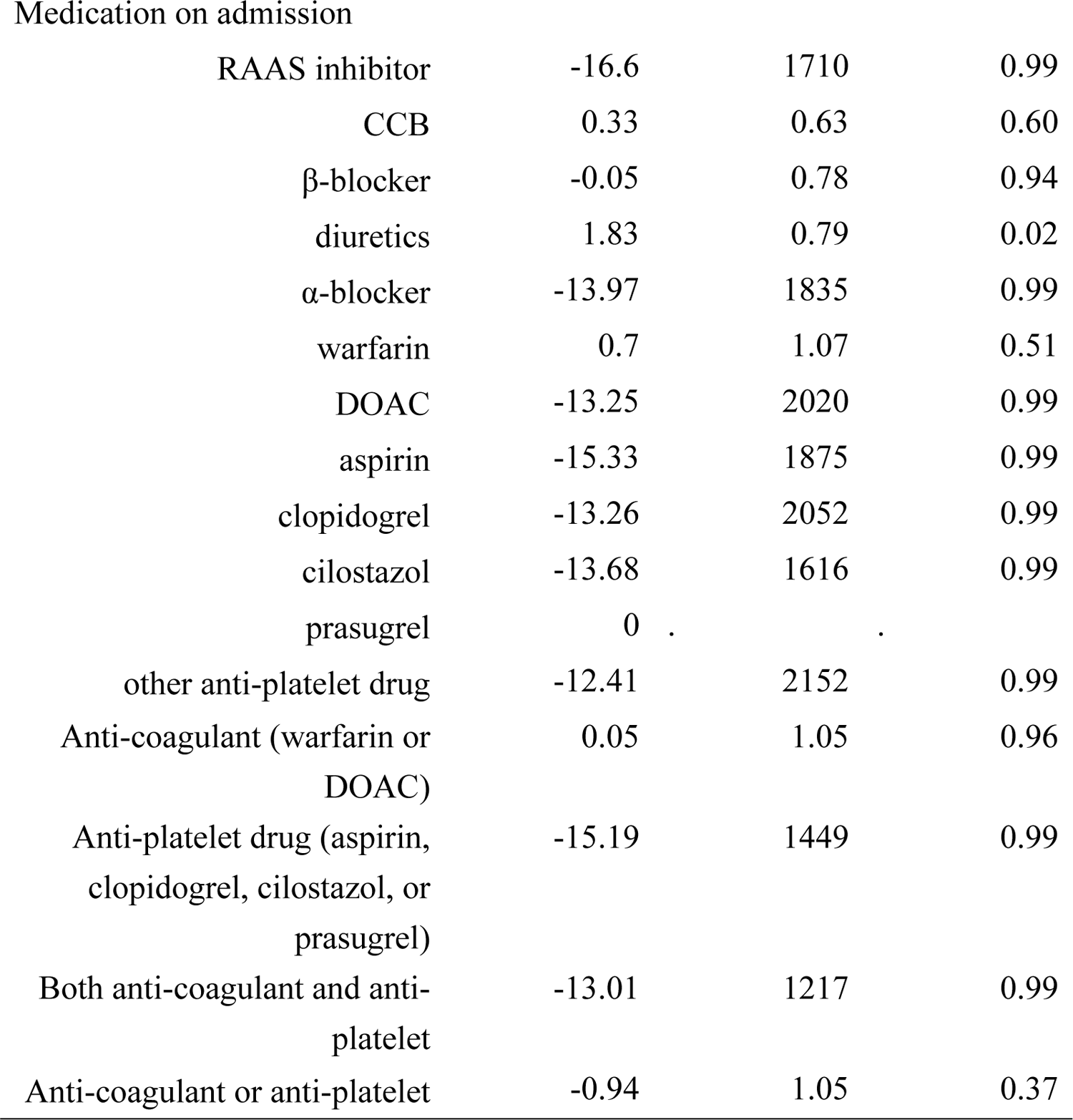
Age and sex-adjusted Cox proportional hazard model in transferred acute **aortic dissection patients (type B)**

| | $\beta$ | SE | p-value |
| --- | --- | --- | --- |
| Systolic BP | -0.03 | 0.01 | 0.01 |
| Diastolic BP | -0.02 | 0.02 | 0.18 |
| Heart rate | 0.03 | 0.02 | 0.05 |
| eGFR | -0.02 | 0.01 | 0.02 |
| Past history |  |  |  |
| A Fib | 2.05 | 0.69 | <0.01 |
| CAD | -14 | 1480 | 0.99 |
| Other CVD | 0.16 | 0.78 | 0.83 |
| PAD | 0.5 | 0.61 | 0.41 |
| VTE | 0 | . | . |
| LV dysfunction | 0 | . | . |
| HT | 14.07 | 1400 | 0.99 |
| DM | 0.17 | 0.77 | 0.82 |
| DLp | -0.02 | 0.66 | 0.98 |
| Genetic and others | -14.11 | 1513 | 0.99 |
| Smoking | -0.01 | 0.7 | 0.99 |
| Alcohol | -0.39 | 0.6 | 0.52 |
| post-AVR | -14.02 | 1587 | 0.99 |
| post-MVR | -11.13 | 1515 | 0.99 |
| CABG | -12.16 | 1558 | 0.99 |
| Intervention to aortic<br>aneurysm and/or dissection | -0.41 | 1.05 | 0.70 |
| Complication |  |  |  |
| Major bleeding | 4.24 | 0.71 | <0.01 |
| Infarction by aortic dissection | 2.04 | 0.57 | <0.01 |
| Paroxymal A Fib | -13.95 | 1435 | 0.99 |
| Medication on admission |  |  |  |
| RAAS inhibitor | -16.6 | 1710 | 0.99 |
| CCB | 0.33 | 0.63 | 0.60 |
| β-blocker | -0.05 | 0.78 | 0.94 |
| diuretics | 1.83 | 0.79 | 0.02 |
| α-blocker | -13.97 | 1835 | 0.99 |
| warfarin | 0.7 | 1.07 | 0.51 |
| DOAC | -13.25 | 2020 | 0.99 |
| aspirin | -15.33 | 1875 | 0.99 |
| clopidogrel | -13.26 | 2052 | 0.99 |
| cilostazol | -13.68 | 1616 | 0.99 |
| prasugrel | 0 |  |  |
| other anti-platelet drug | -12.41 | 2152 | 0.99 |
| Anti-coagulant (warfarin or DOAC) | 0.05 | 1.05 | 0.96 |
| Anti-platelet drug (aspirin, clopidogrel, cilostazol, or prasugrel) | -15.19 | 1449 | 0.99 |
| Both anti-coagulant and anti-platelet | -13.01 | 1217 | 0.99 |
| Anti-coagulant or anti-platelet | -0.94 | 1.05 | 0.37 |

### Cox proportional hazards regression analysis of all-cause death in HOSPITALIZED patients in type A

In the analysis of all-cause death, systolic blood pressure, eGFR, complication of surgical procedures, and use of α-blocker and anti-thrombosis drugs were significantly associated with all-cause death (**Table 7**). The use of anti-thrombosis (warfarin, DOAC, aspirin, clopidogrel) significantly reduced hazard ratio (**Table 7**).

**Table 7.** Cox proportional hazard model in hospitalized acute aortic dissection **patients (type A)**

| | $\beta$ | SE | p-value |
| --- | --- | --- | --- |
| Age | 0.006 | 0.01 | 0.65 |
| Sex | -0.11 | 0.31 | 0.72 |
| Systolic BP | -0.02 | 0.006 | <0.01 |
| Diastolic BP | -0.005 | 0.01 | 0.63 |
| Heart rate | 0.01 | 0.008 | 0.11 |
| eGFR | -0.028 | 0.007 | <0.01 |
| Past history |  |  |  |
| A Fib | -1.34 | 1.01 | 0.19 |
| CAD | 0.67 | 0.53 | 0.21 |
| Other CVD | 0.22 | 0.42 | 0.60 |
| PAD | 0.04 | 0.45 | 0.93 |
| VTE | -12.01 | 915.23 | 0.99 |
| LV dysfunction | -13.04 | 930.19 | 0.99 |
| HT | -0.28 | 0.48 | 0.56 |
| DM | -0.56 | 0.73 | 0.44 |
| DLp | -0.23 | 0.42 | 0.58 |
| Genetic and others | 0.47 | 0.77 | 0.54 |
| Smoking | -0.52 | 0.36 | 0.15 |
| Alcohol | -0.52 | 0.38 | 0.18 |
| post-AVR | 1.03 | 0.73 | 0.16 |
| post-MVR | 0 |  |  |
| CABG | -12.01 | 776.4 | 0.99 |
| Intervention to aortic<br>aneurysm and/or dissection | 0.1 | 0.61 | 0.87 |
| Complication |  |  |  |
| Major bleeding | 1.66 | 0.38 | <0.01 |
| Infarction by aortic dissection | 1.63 | 0.33 | <0.01 |
| Paroxymal A Fib | -1.27 | 0.53 | 0.02 |
| Medication on admission |  |  |  |
| RAAS inhibitor | -0.073 | 0.38 | 0.85 |
| CCB | 0.54 | 0.35 | 0.12 |
| β-blocker | -0.1 | 0.49 | 0.84 |
| diuretics | 0.58 | 0.54 | 0.28 |
| α-blocker | 1.32 | 0.61 | 0.03 |
| warfarin | 0.23 | 0.61 | 0.71 |
| DOAC | -13.03 | 969.9 | 0.99 |
| aspirin | 0.61 | 0.54 | 0.26 |
| clopidogrel | -14.21 | 884.34 | 0.99 |
| cilostazol | -13.04 | 823.72 | 0.99 |
| prasugrel | 0 | . | . |
| other anti-platelet drug | 1.69 | 1.02 | 0.10 |
| Anti-thrombus during hospitalization |  |  |  |
| Anti-coagulant (warfarin or DOAC) | -1.62 | 0.6 | <0.01 |
| Anti-platelet drug (aspirin, clopidogrel, cilostazol, or prasugrel) | -2.88 | 1.01 | <0.01 |
| Both anti-coagulant and anti-platelet | -15.2 | 860.56 | 0.99 |
| Anti-coagulant or anti-platelet | -2.12 | 0.53 | <0.01 |

After adjustment for age and sex, systolic blood pressure, eGFR, complication of surgical procedures, and use of α-blocker were significantly associated with all-cause death (**Table 8**). Also in this analysis, the use of anti-thrombosis was also the significant and strong predictor of reduced all cause death (**Table 8**).

**Table 8.** Age and sex-adjusted Cox proportional hazard model in hospitalized acute **aortic dissection patients (type A)**

| | $\beta$ | SE | p-value |
| --- | --- | --- | --- |
| Systolic BP | -0.02 | 0.006 | <0.01 |
| Diastolic BP | -0.005 | 0.01 | 0.61 |
| Heart rate | 0.012 | 0.008 | 0.12 |
| eGFR | -0.028 | 0.007 | <0.01 |
| Past history |  |  |  |
| A Fib | -1.48 | 1.02 | 0.15 |
| CAD | 0.63 | 0.53 | 0.23 |
| Other CVD | 0.17 | 0.42 | 0.69 |
| PAD | -0.04 | 0.46 | 0.93 |
| VTE | -12.04 | 919.33 | 0.99 |
| LV dysfunction | -13.02 | 930.55 | 0.99 |
| HT | -0.33 | 0.48 | 0.50 |
| DM | -0.58 | 0.73 | 0.42 |
| DLp | -0.2 | 0.42 | 0.63 |
| Genetic and others | 0.58 | 0.78 | 0.46 |
| Smoking | -0.78 | 0.42 | 0.06 |
| Alcohol | -0.65 | 0.43 | 0.13 |
| post-AVR | 1.004 | 0.75 | 0.18 |
| post-MVR | 0 |  |  |
| CABG | -12.14 | 778.45 | 0.99 |
| Intervention to aortic aneurysm and/or dissection | 0.019 | 0.63 | 0.98 |
| Complication |  |  |  |
| Major bleeding | 1.62 | 0.39 | <0.01 |
| Infarction by aortic dissection | 1.64 | 0.33 | <0.01 |
| Paroxymal A Fib | -1.34 | 0.53 | 0.01 |
| Medication on admission |  |  |  |
| RAAS inhibitor | -0.11 | 0.38 | 0.77 |
| CCB | 0.52 | 0.36 | 0.14 |
| β-blocker | -0.15 | 0.49 | 0.76 |
| diuretics | 0.52 | 0.54 | 0.34 |
| α-blocker | 1.31 | 0.61 | 0.03 |
| warfarin | 0.14 | 0.63 | 0.83 |
| DOAC | -13.12 | 972.53 | 0.99 |
| aspirin | 0.55 | 0.55 | 0.31 |
| clopidogrel | -14.39 | 881.58 | 0.99 |
| cilostazol | -13.05 | 824.28 | 0.99 |
| prasugrel | 0 | . | . |
| other anti-platelet drug | 1.62 | 1.03 | 0.12 |
| Anti-thrombus during hospitalization |  |  |  |
| Anti-coagulant (warfarin or DOAC) | -1.64 | 0.6 | <0.01 |
| Anti-platelet drug (aspirin, clopidogrel, cilostazol, or prasugrel) | -2.9 | 1.01 | <0.01 |
| Both anti-coagulant and anti-platelet | -15.19 | 859.87 | 0.99 |
| Anti-coagulant or anti-platelet | -2.17 | 0.53 | <0.01 |

### Cox proportional hazards regression analysis of all-cause death in HOSPITALIZED patients in type B

In the analysis of all-cause death, heart rate, history of atrial fibrillation and complication of surgical procedures were significantly associated with all-cause death (**Table 9**). In this analysis, the use of anti-thrombosis was not the predictor of all cause death predictor of all cause death (**Table 9**).

**Table 9.**
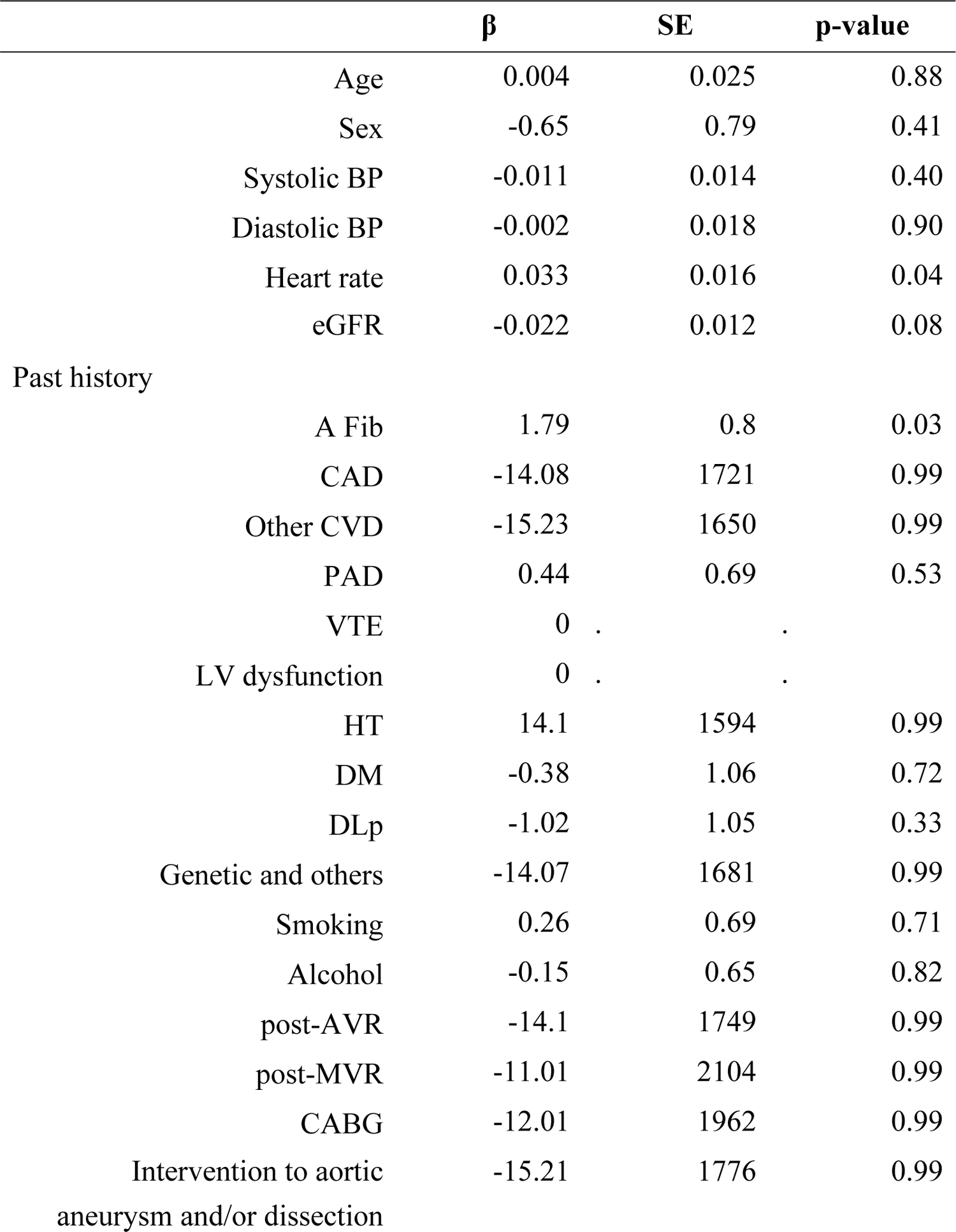
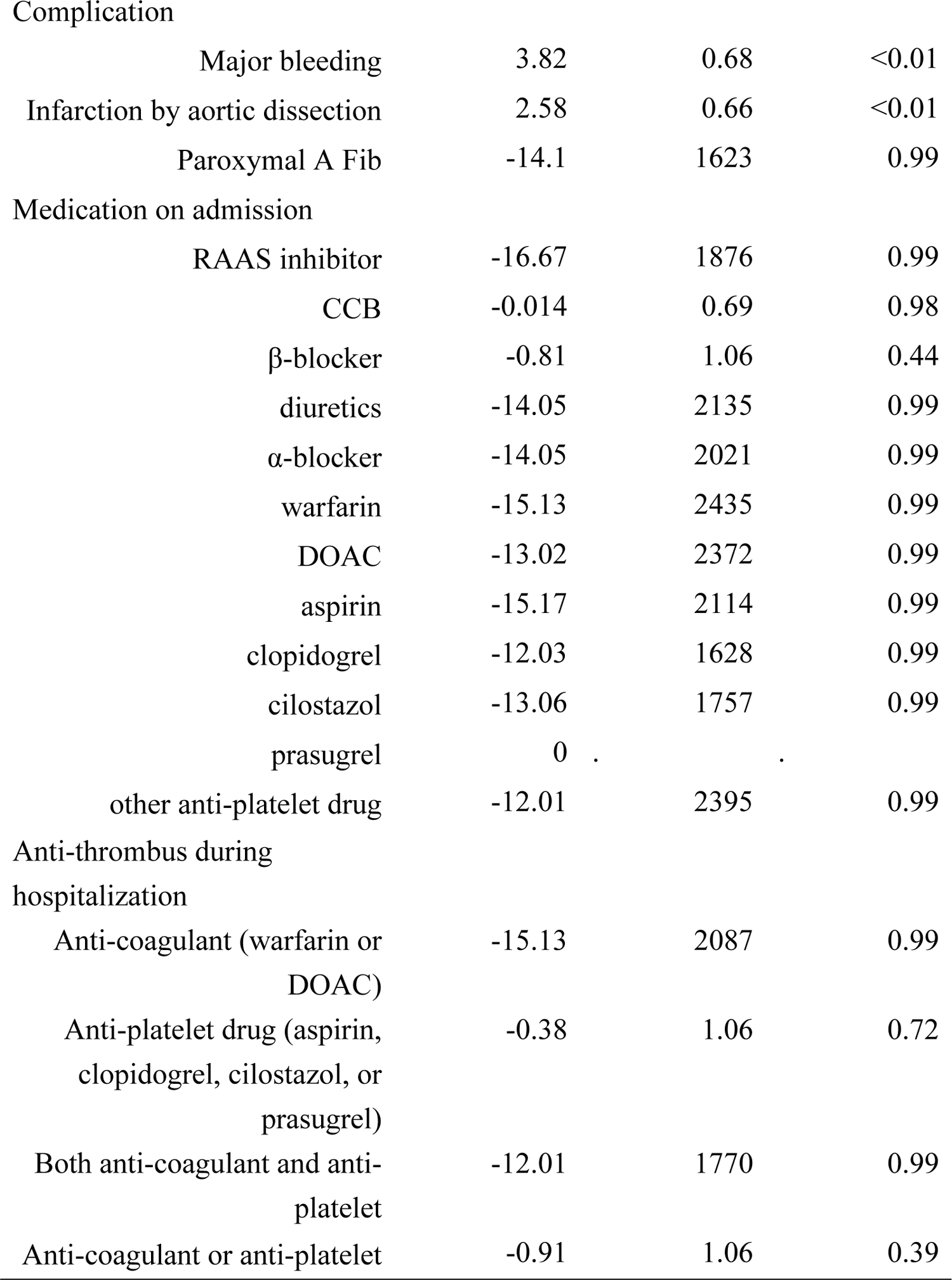
Cox proportional hazard model in hospitalized acute aortic dissection patients (type B)

| | $\beta$ | SE | p-value |
| --- | --- | --- | --- |
| Age | 0.004 | 0.025 | 0.88 |
| Sex | -0.65 | 0.79 | 0.41 |
| Systolic BP | -0.011 | 0.014 | 0.40 |
| Diastolic BP | -0.002 | 0.018 | 0.90 |
| Heart rate | 0.033 | 0.016 | 0.04 |
| eGFR | -0.022 | 0.012 | 0.08 |
| Past history |  |  |  |
| A Fib | 1.79 | 0.8 | 0.03 |
| CAD | -14.08 | 1721 | 0.99 |
| Other CVD | -15.23 | 1650 | 0.99 |
| PAD | 0.44 | 0.69 | 0.53 |
| VTE | 0 | . | . |
| LV dysfunction | 0 | . | . |
| HT | 14.1 | 1594 | 0.99 |
| DM | -0.38 | 1.06 | 0.72 |
| DLp | -1.02 | 1.05 | 0.33 |
| Genetic and others | -14.07 | 1681 | 0.99 |
| Smoking | 0.26 | 0.69 | 0.71 |
| Alcohol | -0.15 | 0.65 | 0.82 |
| post-AVR | -14.1 | 1749 | 0.99 |
| post-MVR | -11.01 | 2104 | 0.99 |
| CABG | -12.01 | 1962 | 0.99 |
| Intervention to aortic aneurysm and/or dissection | -15.21 | 1776 | 0.99 |

|  |  |  |  |  |
| --- | --- | --- | --- | --- |
| Complication |  |  |  |  |
|  | Major bleeding | 3.82 | 0.68 | <0.01 |
|  | Infarction by aortic dissection | 2.58 | 0.66 | <0.01 |
|  | Paroxymal A Fib | -14.1 | 1623 | 0.99 |
| Medication on admission |  |  |  |  |
|  | RAAS inhibitor | -16.67 | 1876 | 0.99 |
|  | CCB | -0.014 | 0.69 | 0.98 |
| | β-blocker | -0.81 | 1.06 | 0.44 |
|  | diuretics | -14.05 | 2135 | 0.99 |
| | α-blocker | -14.05 | 2021 | 0.99 |
|  | warfarin | -15.13 | 2435 | 0.99 |
|  | DOAC | -13.02 | 2372 | 0.99 |
|  | aspirin | -15.17 | 2114 | 0.99 |
|  | clopidogrel | -12.03 | 1628 | 0.99 |
|  | cilostazol | -13.06 | 1757 | 0.99 |
|  | prasugrel | 0 |  |  |
|  | other anti-platelet drug | -12.01 | 2395 | 0.99 |
| Anti-thrombus during hospitalization |  |  |  |  |
|  | Anti-coagulant (warfarin or DOAC) | -15.13 | 2087 | 0.99 |
|  | Anti-platelet drug (aspirin, clopidogrel, cilostazol, or prasugrel) | -0.38 | 1.06 | 0.72 |
|  | Both anti-coagulant and anti-platelet | -12.01 | 1770 | 0.99 |
|  | Anti-coagulant or anti-platelet | -0.91 | 1.06 | 0.39 |

After adjustment for age and sex, history of atrial fibrillation and complication of surgical procedures were significantly associated with all-cause death (**Table 10**). Also, the use of anti-thrombosis was not the predictor of all cause death predictor of all cause death (**Table 10**).

**Table 10.** Age and sex-adjusted Cox proportional hazard model in hospitalized acute aortic dissection patients (type B)

| | $\beta$ | SE | p-value |
| --- | --- | --- | --- |
| Systolic BP | -0.012 | 0.014 | 0.37 |
| Diastolic BP | -0.005 | 0.019 | 0.79 |
| Heart rate | 0.035 | 0.018 | 0.05 |
| eGFR | -0.025 | 0.013 | 0.05 |
| Past history |  |  |  |
| A Fib | 1.87 | 0.83 | 0.02 |
| CAD | -13.99 | 1722 | 0.99 |
| Other CVD | -15.32 | 1649 | 0.99 |
| PAD | 0.44 | 0.71 | 0.53 |
| VTE | 0 | . | . |
| LV dysfunction | 0 | . | . |
| HT | 14.09 | 1589 | 0.99 |
| DM | -0.32 | 1.06 | 0.76 |
| DLp | -1 | 1.06 | 0.34 |
| Genetic and others | -13.84 | 1682 | 0.99 |
| Smoking | -0.099 | 0.8 | 0.90 |
| Alcohol | -0.42 | 0.68 | 0.54 |
| post-AVR | -13.79 | 1754 | 0.99 |
| post-MVR | -11.24 | 2140 | 0.99 |
| CABG | -12.21 | 1995 | 0.99 |
| Intervention to aortic<br>aneurysm and/or dissection | -15.16 | 1769 | 0.99 |
| Complication |  |  |  |
| Major bleeding | 4.17 | 0.81 | <0.01 |
| Infarction by aortic dissection | 2.6 | 0.66 | <0.01 |
| Paroxymal A Fib | -13.95 | 1622 | 0.99 |
| Medication on admission |  |  |  |
| RAAS inhibitor | -16.64 | 1879 | 0.99 |
| CCB | 0.14 | 0.71 | 0.85 |
| β-blocker | -0.7 | 1.02 | 0.51 |
| diuretics | -14.13 | 2151 | 0.99 |
| α-blocker | -13.99 | 2021 | 0.99 |
| warfarin | -14.86 | 2386 | 0.99 |
| DOAC | -13.32 | 2347 | 0.99 |
| aspirin | -15.46 | 2096 | 0.99 |
| clopidogrel | -12.25 | 1637 | 0.99 |
| cilostazol | -13.7 | 1797 | 0.99 |
| prasugrel | 0 |  |  |
| other anti-platelet drug | -12.37 | 2454 | 0.99 |
| Anti-thrombus during hospitalization |  |  |  |
| Anti-coagulant (warfarin or DOAC) | -15.03 | 2073 | 0.99 |
| Anti-platelet drug (aspirin, clopidogrel, cilostazol, or prasugrel) | -0.31 | 1.08 | 0.78 |
| Both anti-coagulant and anti-platelet | -12.29 | 1802 | 0.99 |
| Anti-coagulant or anti-platelet | -0.82 | 1.08 | 0.44 |

## Discussion

This is the first report to demonstrate that anti-thrombotic drugs did not worsen the prognosis in patients with acute aortic dissection, in whom higher prevalence of previous history of atrial fibrillation and cardiovascular diseases were observed. Although there might be some hidden better conditions, the use of anti-thrombotic drugs during hospitalization showed significantly better prognosis in type A, which was observed even after adjustment for age and sex. Probably because more attention is paid in surgical repair in patients with anti-thrombotic drugs, anti-thrombotic drugs did not worsen the prognosis in acute aortic dissection. It is unknown if anti-thrombotic drugs have pharmacologically beneficial effects in the clinical course of acute aortic dissection.

### Thrombus in False Lumen in Acute Aortic Dissection

After aortic dissection development, thrombus is often created in false lumen. It has been reported that the false lumen status is associated with poor outcomes[12]. The meta-analysis indicated that partial thrombosis in false lumen is independently associated with poor long-term survival in acute aortic dissection, compared with complete thrombosis or patent false lumen[12]. Although the underlying mechanisms involved in the association between false lumen status and outcomes are not fully elucidated, complete thrombus formation of the false lumen has been considered to be a prerequisite for healing the aorta[12]. Also, complete thrombus may be able to relieve pressure from the tear; however, it may cause organ ischemia. In partial thrombosis in false lumen, organ ischemia may occur, and further, thrombus and platelets can modulate inflammatory reactions and immune responses. [13] [14] Although it remains unknown how anti-thrombotic drugs work on the thrombus formation in false lumen, the present study indicated that anti-thrombotic drugs did not worsen the prognosis in patients with acute aortic dissection. The mechanisms how anti-thrombotic drugs work in acute aortic dissection should be clarified in near future.

### Initial Medical Therapy in Acute Aortic Dissection

Initial medical therapy should stabilize patients, control pain, and lower blood pressure to prevent further propagation of the dissection and to lessen the risk of aortic rupture.[9] The present study demonstrated that anti-thrombotic drugs do not worsen the prognosis; however, it is not recommended to newly introduce anti-thrombotic drugs before transfer of patients to comprehensive aortic centers or appropriate surgical consultation.[9]

### Medical Management during Hospitalization

Blood pressure management is the most important.[9] β-Adrenergic blockade reduces not only heart rate and blood pressure, but also provides anti-impulse therapy to decrease aortic wall stress.[15] Besides appropriate blood pressure management, other therapeutic medical strategies are required. Although the present study demonstrated that the prognosis was comparable between with and without anti-thrombotic drugs during hospitalization in type B aortic dissection, anti-thrombotic drug group showed significantly better prognosis in patients with type A aortic dissection, suggesting that anti-thrombotic drugs might be a new therapeutic strategy in type A. On the other hand, anti-thrombotic drugs might not have been administered in poor condition, even if they were indicated. In the present study, we were not able to exclude confounding by indication.

### Long-term Medical Management

Survival rate has been reported between 52% and 94% at 1 year and between 45% and 88% at 5 years in type A,[16] and between 56% and 92 % at 1 year and between 48% and 82% at 5 years in type B.[17] Long-term management of acute aortic dissection should be performed to prevent late phase complications, including aneurysmal enlargement and aortic rupture. Poorly controlled hypertension has been reported to increase late morbidity and mortality.[18, 19] Thus, it is required to perform blood pressure control, screening the patient and relatives for heritable disorders associated with aortic dissection, serial imaging of the aorta, lifestyle modifications, and education.[16, 20] It should be clarified if anti-thrombotic drugs improve in these patients in near future.

### Limitations

The present study has several limitations. First, the present study was an observational retrospective cohort study from a single center. Anti-thrombotic drugs were not administered for aortic dissection, but for other cardiovascular diseases, such as coronary artery diseases or atrial fibrillation. Second, the enrolled patients in the present study were hospitalized in University Hospital. These patients might be in more severe condition. Third, this study was not able to include the hidden severity data of acute aortic dissection except the obtained medical records in the present study, which may affect the prognosis. Fourth, confounding by indication cannot be excluded in anti-thrombotic drugs use in hospitalized patients with type A aortic dissection. Fifth, we have no available data of long-term prognosis.

## Conclusion

We demonstrated that anti-thrombotic drugs did not worsen the prognosis in patients with acute aortic dissection, indicating that we should not hesitate anti-thrombotic drugs if indicated.

## Data Availability

All relevant data are within the manuscript and its Supporting Information files.

## Acknowledgements

None

## References

1. Goebel N, Nagib R, Salehi-Gilani S, Ahad S, Albert M, Ursulescu A, et al. One-stage hybrid aortic repair using the frozen elephant trunk in acute DeBakey type I aortic dissection. J Thorac Dis. 2018;10:4195–203.

2. Shimamoto T, Komiya T, Tsuneyoshi H. Fate of uncomplicated acute type B aortic dissection and impact of concurrent aortic dilatation on remote aortic events. J Thorac Cardiovasc Surg. 2019;157:854–863.

3. Nienaber CA, Eagle KA. Aortic dissection: new frontiers in diagnosis and management: Part I: from etiology to diagnostic strategies. Circulation. 2003;108:628–35.

4. Hirst AE, Jr., Johns VJ, Jr., Kime SW, Jr. Dissecting aneurysm of the aorta: a review of 505 cases. Medicine (Baltimore). 1958;37:217–279.

5. Fukui T. Management of acute aortic dissection and thoracic aortic rupture. J Intensive Care. 2018;6:15.

6. Kimura T, Shiraishi K, Furusho A, Ito S, Hirakata S, Nishida N, et al. Tenascin C protects aorta from acute dissection in mice. Sci Rep. 2014;4:4051.

7. Tahara N, Hirakata S, Okabe K, Tahara A, Honda A, Igata S, et al. FDG-PET/CT images during 5 years before acute aortic dissection. Eur Heart J. 2016;37:1933.

8. Ohno-Urabe S, Aoki H, Nishihara M, Furusho A, Hirakata S, Nishida N, et al. Role of Macrophage Socs3 in the Pathogenesis of Aortic Dissection. J Am Heart Assoc. 2018;7:e007389.

9. Malaisrie SC, Szeto WY, Halas M, Girardi LN, Coselli JS, Sundt TM, 3rd, et al. 2021 The American Association for Thoracic Surgery expert consensus document: Surgical treatment of acute type A aortic dissection. J Thorac Cardiovasc Surg. 2021;162:735-758 e2.

10. Schulman S, Kearon C, Subcommittee on Control of Anticoagulation of the S, Standardization Committee of the International Society on T, Haemostasis. Definition of major bleeding in clinical investigations of antihemostatic medicinal products in non-surgical patients. J Thromb Haemost. 2005;3:692–694.

11. Matsuo S, Imai E, Horio M, Yasuda Y, Tomita K, Nitta K, et al. Revised equations for estimated GFR from serum creatinine in Japan. Am J Kidney Dis. 2009;53:982–992.

12. Li D, Ye L, He Y, Cao X, Liu J, Zhong W, et al. False Lumen Status in Patients With Acute Aortic Dissection: A Systematic Review and Meta-Analysis. J Am Heart Assoc. 2016;5:e003172.

13. Aoki H, Majima R, Hashimoto Y, Hirakata S, Ohno-Urabe S. Ying and Yang of Stat3 in pathogenesis of aortic dissection. J Cardiol. 2021;77:471–474.

14. Hamad MA, Krauel K, Schanze N, Gauchel N, Stachon P, Nuehrenberg T, et al. Platelet Subtypes in Inflammatory Settings. Front Cardiovasc Med. 2022;9:823549.

15. Svensson LG, Crawford ES. Aortic dissection and aortic aneurysm surgery: clinical observations, experimental investigations, and statistical analyses. Part I. Curr Probl Surg. 1992;29:817–911.

16. Braverman AC. Acute aortic dissection: clinician update. Circulation. 2010;122:184–188.

17. Tsai TT, Fattori R, Trimarchi S, Isselbacher E, Myrmel T, Evangelista A, et al. Long-term survival in patients presenting with type B acute aortic dissection: insights from the International Registry of Acute Aortic Dissection. Circulation. 2006;114:2226–2231.

18. Melby SJ, Zierer A, Damiano RJ, Jr., Moon MR. Importance of blood pressure control after repair of acute type a aortic dissection: 25-year follow-up in 252 patients. J Clin Hypertens (Greenwich). 2013;15:63–68.

19. Chan KK, Lai P, Wright JM. First-line beta-blockers versus other antihypertensive medications for chronic type B aortic dissection. Cochrane Database Syst Rev. 2014;:CD010426.

20. Chaddha A, Eagle KA, Braverman AC, Kline-Rogers E, Hirsch AT, Brook R, et al. Exercise and Physical Activity for the Post-Aortic Dissection Patient: The Clinician’s Conundrum. Clin Cardiol. 2015;38:647–651.

